# Decoding molecular programs in melanoma brain metastases

**DOI:** 10.1101/2022.02.06.22270509

**Authors:** Josefine Radke, Elisa Schumann, Julia Onken, Randi Koll, Güliz Acker, Bohdan Bodnar, Carolin Senger, Sascha Tierling, Markus Möbs, Peter Vajkoczy, Anna Vidal, Sandra Högler, Petra Kodajova, Dana Westphal, Friedegund Meier, Frank Heppner, Susanne Kreuzer-Redmer, Florian Grebien, Karsten Jürchott, Torben Redmer

## Abstract

The systemic dissemination of tumor cells and the spatiotemporal development of organ- metastases are associated with loss of therapeutic control and decreased overall survival (OS). The emergence of solitary or multiple brain metastases is frequently observed in melanoma patients and responsible for disease progression and dismal prognosis. Here, we used whole transcriptome and methylome profiling as well as targeted sequencing (TargetSeq) of intraoperative/snap frozen or archived melanoma brain metastases to unravel molecular subgroups and subclonal heterogeneity. We discovered that E-cadherin (Ecad)/BRAF^V600E/K^, CD271/NRAS^Q61L/R/K^, and tumor infiltrated lymphocytes (TIL)-status molecularly subdivided tumors into proliferative/pigmented and invasive/stem-like irrespective of the intracranial location. Moreover, we identified 46 differentially methylated regions in promoters of 14 genes, subdividing MBM into BRAF^mut^ and NRAS^mut^ subgroups. We observed that therapy-resistant, migratory CD271^+^/Ecad^neg^ subclones derived from Ecad^+^tumors in an epithelial-mesenchymal transition (EMT)-like process and fostered intracranial progression. Hence, CD271^high^ MBM present a therapy-resistant, progressive subset of tumors that are refractory to conventional therapeutic strategies. The knockdown of CD271 or SOX4 in *in vitro* established, MBM-derived cell lines decreased cell migration, proliferation, and number of suspension cells that were shed by cell lines of progressive tumors. In summary, we propose that an Ecad-to-CD271 switch of MBM is a rate-limiting process that potentially determines intracranial progression in melanoma patients. The therapeutic control of this process may prevent intracranial progression, increasing patient’s overall survival.

## Introduction

The development of brain metastases (BM) is frequent in melanoma, lung and breast cancer^1, 2^. Despite much progress and remarkable response in a subset of patients^3^, small molecule inhibitors (BRAFi) or immune checkpoint inhibitors (ICi) blocking oncogenic BRAF or interfering with the PD-L1/PD1 axis to restore T cell activation are insufficient strategies to achieve long-lasting prevention of intracranial relapse and progression^4, 5^. The latter is determined by emergence of multiple BM and therefore associated with poor prognosis^3, 6^.

Stage IV melanoma patients exhibit extracranial metastases at multiple organ sites facilitated by circulating tumor cells^7^ invading the sub-arachnoid space and cerebrospinal fluid (CSF)^8^. It is assumed that primary tumor-derived cells acquire the capability of transmigration through the blood brain barrier (BBB) during the sequential steps of the metastatic cascade. Due to the difficulty of tracking of cellular subclones giving rise to BM human patients are seldom accessible to investigation. Therefore, the investigation of single BM, each presenting temporal snapshots, provides insights into the mechanisms of BM development and progression.

Melanoma cells that crossed the BBB initially remain in a dormant state and likely develop symptomatic macrometastases after a certain time of adaptation to the brain microenvironment or environmental trigger^9–11^. In fact, brain metastatic lesions are observed in more than 90% of melanoma patients *post mortem*^12, 13^, proposing that only a minority of micrometastases successfully establishes macrometastases. The time from initial diagnosis of primary tumors to the detection of BM ranges from 1-10 years, supporting the assumption of a slow evolutionary process of BM in 20 – 40% of melanoma patients^3, 14^. The median overall survival of melanoma patients after diagnosis of a BM (MBM) is 8.9 months^15^ and is determined by the speed of intracranial progression that in turn depends on the efficacy of development of macrometastases and the response to therapeutic interventions.

MBM like extracranial metastases (Met) comprise numerous genetic subclones that variably respond to BRAFi, ICi and radiotherapy due to intrinsic or acquired mechanisms of resistance consequently fostering tumor relapse within approximately 6-11 months^16–18^. So far, the presence or acquisition of mutations in NRAS (p.Q61K/L), MEK1 (p.P124), RAC1 (p.P29S) or gain of a secondary mutation in BRAF (p.L514K) have been attributed to confer BRAFi resistance^19, 20^. In addition, BM exhibiting increased DNA repair capacity and/or low proliferative capacity present a low response to radiation therapy^21^ and melanoma expressing IPRES (innate anti-PD-1 resistance) signature genes is resistant to ICi-based therapies^22^. However, expression of cell surface markers serving as targets of antibody-based therapies is unstable and regulated by phenotype switching. The latter process is responsible for a non-genomic cellular heterogeneity and is triggered by pro-inflammatory cytokines that are released in response to therapy-induced tumor cell stress or from microenvironmental cells such as astrocytes or microglia^23, 24^. In particular, astrocytes are implicated in the maintenance of BBB permeability, extracellular homeostasis and essentially involved in the response towards brain damaging events^25, 26^. The interaction of brain tumors with astrocytes triggers astrogliosis, that results in secretion of cytokines promoting the survival and invasion of tumor cells^27^. Hence, inflammation-triggered mechanisms might drive the progression of MBM via a subset of dedifferentiated and highly migratory cancer stem-like tumor cells (CSCs)^23, 28^. The latter comprise a minor tumor cell subset exhibiting cellular plasticity, display elevated intrinsic resistance to several therapeutic drugs and feature a neural crest stem cell (NCSC)-like phenotype in melanoma^29–31^. The maintenance of the NCSC-state of melanoma cells depends on the expression of the nerve growth factor receptor and non-receptor tyrosine kinase CD271 that is associated with a network of several downstream targets^28, 32–34^ and non-genetic emergence of minimal-residual disease (MRD)^28, 33–36^. Recent work has shown that melanoma cells secreted extracellular vesicles were enriched with CD271 that was taken up by lymphatic endothelial cells, therefore aiding lymph node metastasis^37^.

On the other hand, CD271 controls migratory programs of melanocytes^38^ that are connected with keratinocytes via E-cadherin (Ecad)-mediated adhesive junctions in the skin. The downregulation of Ecad is tightly controlled by a set of transcription factors mediating the transition of epithelial to mesenchymal (EMT) states and is a prerequisite for melanocyte migration and their malignant transformation^39^. However, the expression of Ecad is essential to establish stemness and is restored in primary melanoma and organ-specific metastases^40–42^.

Likely, BM emerge and progress by the concerted interaction of several molecular programs that are triggered by cells of the tumor microenvironment (TME) and/or in response to therapeutic interventions. To gain insight into molecular mechanisms controlling the maintenance and progression of BM, we performed a whole transcriptome and methylome profiling as well as targeted sequencing (TargetSeq) of intraoperative or cryo-preserved MBM (n=16) and established MBM-derived cellular model systems. Here, we provided evidence that therapeutic interventions inhibiting the acquired constitutive activation of BRAF likely foster the phenotype switch from Ecad^+^ into NGFR/CD271^+^ cells.

## Material and Methods

### Patient cohorts

Intraoperative brain metastases of sixteen patients with diagnosed stage IV melanoma (MBM) were surgically removed at the Department of Neurosurgery, Charité – Universitätsmedizin Berlin, Germany. Tumor pieces were split into parts of equal size and either i) snap-frozen and stored at -80 °C or ii) directly used for the establishment of MBM-derived cell lines or iii) isolation of RNA and DNA or formaldehyde-fixed and paraffin-embedded (FFPE) and archived. Cerebrospinal fluid (CSF) of patient 8 was collected by lumbar puncture. All patients gave written informed consent for the collection and scientific use of tumor material which was collected at the Biobank of the Charité – Comprehensive Cancer Center (CCCC) following ethics approval (EA1/152/10; EA1/107/17; EA4/028/18). In addition, thirty-two MBM archived at the Department of Neuropathology, Charité-Universitätsmedizin Berlin, Germany, were included in the study and analyzed. The usage of archived (FFPE) melanoma and central nervous system-derived control samples (pons, cortex, cerebellum) has been reviewed and approved by the local ethics committee (EA1/107/17 and EA1/075/19).

### Cell culture

#### Conventional melanoma cell lines

BMCs and conventional melanoma cell lines were kept at 37°C/ 5% CO_2_ and 95% humidity in cell culture medium (DMEM, 4.5 g/L glucose, stabilized glutamine/GlutaMax, pyruvate, Gibo/ThermoFisher) supplemented with 10% fetal bovine (FBS, Gibco) serum and 1% penicillin/streptomycin (P/S) (Gibo/ThermoFisher). BMCs were kept at low passages (2-20) and split according to their proliferative capacity (1:2-1:10) at a confluence of ∼80 %. Cells were seeded onto glass 8-chamber slides to a density of 5,000-10,000 cells per chamber. A375, T2002, MeWo cells and human melanocytes were cultured as previously reported^33^.

### MBM-derived cell lines

Intraoperative tumors were surgically resected during routine craniotomy and processed to establish brain metastases-derived cell lines (BMCs) as following: Tumor pieces were stored in physiological saline, 0.9% on ice until further processing. Following mincing using scalpels, the mechanically dissociated tissue was transferred to a 15 ml falcon tube containing trypsin/EDTA (0.05%) and incubated at 37°C in a water bath for up to 20 minutes. In addition, the tissue was mechanically dissociated by usage of a Pellet Mixer (VWR International). The cell suspension was applied to a 70 µm cell strainer to remove undigested tissue fragments and cells in the flow-through were collected by centrifugation at 330g for 5 minutes. Collected cells were resuspended in cell culture medium (DMEM, 4.5 g/L glucose, stabilized (GlutaMax) or conventional glutamine, pyruvate, 10% FBS, 1% P/S) and seeded on appropriate cell culture dishes. Cells were maintained for at least three days without medium change to achieve optimal recovering and attachment of tumor cells.

### BrdU labeling

For labeling, cells were maintained for 2h in medium containing BrdU (Becton&Dickinson) at a final concentration of 2 mM. Subsequently, cells were washed with phosphate buffered saline (PBS) and fixed with freshly prepared paraformaldehyde (4% in PBS) for 10 min at room temperature and washed and permeabilized by Triton-X100 (0.1 %/PBS). Following, cells were treated with hydrochloric acid (2M) for 10 min and washed twice with PBS. For BrdU detection, labeled cells were incubated with anti-BrdU-AlexaFluor488 for 1h at room temperature or overnight at 4°C and washed with PBS-Tween20 (PBST; 0.1%/PBS). Images were taken with Leica fluorescence microscope (Zeiss Axioskop 2) and edited by Adobe Photoshop 2020 using the gradation curve and picture size function. Images were adjusted to a resolution of 600 dpi (RGB). BrdU-positive cells were quantified by counting and related to the total number of cells.

### Flow cytometry/Fluorescence-activated cell sorting (FACS)

After removal of medium, cells were washed with PBS and harvested by Trypsin (0.05 % Trypsin/EDTA). Following addition of cell culture medium, cells were collected by centrifugation at 330g at room temperature for 3 (min) and resuspended in 100 µl of ice cold buffer (PBS/0.5 % bovine serum/2 mM EDTA) and stored on ice. Cells were incubated with fluorescently labeled primary antibodies against CD271-PE (Miltenyi) DECMA1-APC (recognizing the N-terminal domain of E-cadherin, Biolegend), and non-labeled antibodies against AXL (Novus biologicals), PD-L1 (BioLegend), c-MET (MET, Cell signaling) or KBA.62 (BioLegend) diluted in buffer according to the manufacturer’s specifications and stored at 4°C for 10 min to achieve proper labeling. Following, cells were washed by addition of buffer, collected by centrifugation and resuspended in 100µl of buffer that contained secondary antibodies (AlexaFluor-488/594/647) and/or DAPI, diluted according to the manufacturer’s specifications. After incubation for 10 min at 4°C and washing, cells were resuspended in 500 µl PBS and analyzed by flow cytometry (Canto II) or fractioned by FACS using a FACSAria™III cell sorter (Becton&Dickinson, BD). FACS-isolated cells were collected in cell culture medium and seeded on appropriate vessels following centrifugation. Data analysis was performed with FlowJo (Ver 10.7.1).

### Immunophenotyping

#### Immunofluorescence (IF)

Two micrometer sections of FFPE tumors were dewaxed and subjected to antigen retrieval with citrate buffer (10 mM, ph=6.0) and heating for 20 min in a steamer. Cooled sections were blocked with blocking buffer (2% BSA/PBS) to reduce unspecific binding. Primary antibodies (p75NTR, Cell signaling/CST, #8238, mAb rabbit, clone D4B3, 1:100; E-cadherin, CST#3195, mAb rabbit, clone 24E10, 1:200; E-cadherin, Santa Cruz, sc-8426, mAb mouse, clone G10, 1:50; E-cadherin-AlexaFluor647, BioLegend, 147308, mAb rat, clone DECMA-1, 1:50; KBA.62, NovusBiologicals, NBP2-45285, mAb mouse, 1:100; GFAP-AlexaFluor594, BioLegend, 644708, mAb mouse, clone 2E1.E9, 1:200; STAT5A, Abcam, ab32043, mAb rabbit, clone E289, 1:100; AXL, CST, #8661, mAb rabbit, clone C89E7, 1:100 and phospho(Tyr779)-AXL, R&D Systems, mAb mouse, clone 713610, 1:50 were diluted in blocking buffer and incubated for 2h at room temperature or overnight at 4°C. After washing with PBST, secondary antibodies and DAPI all diluted to 1:500 in blocking buffer were applied to sections and incubated at room temperature for 1h. Following washing, sections were covered with mounting medium and cover slips and stored at 4°C until fluorescence microscopy-based imaging.

#### Immunohistochemistry (IHC)

Automated histological staining was performed on the BenchMark Ultra platform (Ventana) or autostainer (Agilent) using p75NTR, CST, #8238, mAb rabbit, clone D4B3, 1:100, c-MET/MET, CST, #8741, mAb mouse, clone L6E7, 1:100), phospho(Tyr1234/1235)-MET CST, #3077, mAb rabbit, clone D26, 1:100; AXL, CST, #8661, mAb rabbit, clone C89E7, 1:100 or STAT5A, Abcam, ab32043, mAb rabbit, clone E289, 1:100.

### RNA isolation and sequencing

Isolation of total RNA from snap frozen or intraoperative tumors was performed with the RNAeasy extraction kit (Quiagen) according to the manufacturer’s instructions. RNA integrity was determined by automated electrophoresis (4200 TapeStation system, Agilent). The library preparation of 100 ng total RNA was performed with TruSeq Stranded total RNA Sample Preparation-Kit and Ribo Zero Gold (Illumina) and paired-end (2x100 bp) whole transcriptome profiling of RNA with integrity numbers (RIN) ≥7 was performed at Cegat GmbH, Tuebingen (Germany) and sequenced on NovaSeq6000 platform. Illumina bcl2fastq (2.19) was used for demultiplexing of sequenced reads and adapter trimming was performed with Skewer (version 0.2.2)^43^. The information on FASTQ files was obtained using the FastQC program (version 0.11.5-cegat) read out. Raw sequencing data (fastq files) were quality controlled using fastqc (version 0.11.7 - Bioinformatics Group at the Babraham Institute) and further preprocessed with fastp^44^. Reads were aligned to the GRCh38 version of the human genome using TopHat^45^ and counts per gene were calculated by the featureCount-algorithm from the Rsubread package^46^. All further steps of the analysis were done in R. Raw counts of protein-coding genes were normalized using the DESeq2 (https://bioconductor.org/packages/release/ bioc/html/DESeq2.html) package^47^. Differential expression of genes between groups was determined after fitting models of negative binomial distributions to the raw counts. Raw p-values were FDR (false discovery rate)- adjusted for multiple testing and a value below 0.05 for the adjusted p-values were used to determine significant differentially expressed genes. Functional annotation of genes, over representation and gene set enrichment analysis were done using the clusterProfiler package^48^. For visualization of differentially expressed genes and molecular subgroups we used ComplexHeatmap^49^ https://www.bioconductor.org/packages/release/bioc/ html/Complex Heatmap.html).

### Genetic profiling

For amplicon-based targeted DNA-sequencing, 10 - 40 ng of DNA was isolated from stored snap frozen and archived FFPE tumor tissue or from cell lines using the DNeasy Blood & Tissue or the QIAamp DNA FFPE Tissue Kit and used for library preparation. Sequencing of cancer hotspot (CHP2v, ThermoFisher) or TruSight Oncology 500 panel (Illumina) libraries was performed with benchtop sequencers IonProton (Thermo fisher) or NextSeq2000 (Illumina) with 775X mean coverage (Supplementary Table 6). Sequencing results were analyzed with VariantCaller software and validated using databases such as Varsome, COSMIC, and the 1000Genomes project^50^. Data of the latter enabled the separation of single nucleotide polymorphisms (SNPs) from mutations (single nucleotide varaints, SNVs). Copy number analyses of targeted-sequencing reads of multiple sites of a certain gene was performed using binary alignment map (BAM) and BAM index (bai) files and the CNVPanelizer R-script (https://www.bioconductor.org/packages/release/bioc/html/ CNVPanelizer.html). We used OncoPrint for visualization of the genomic alterations (https://jokergoo.github.io/ComplexHeatmap-reference/book/oncoprint.html).

### Production of retroviral and lentiviral particles

For production of lentiviral particles, LentiX cells were seeded to 1x10^4^ cells on a 10 cm dish and transfected after 24h with 4 µg of plasmids either expressing the DOX (doxycycline)- inducible, SOX4 targeting shRNA (GEPIR Sox4.2137, Addgene #101119) or mutant RAC1^P29S^ (pHAGE-RAC1-P29S, Addgene #116650) and 2 µg of pMD2.G (Addgene # 12259, VSV-G envelope) and 1 µg of psPAX2 (#12260) packaging plasmids using 20 µl/1ml Polyethylenimine (PEI, Sigma-Aldrich). Medium was changed after 24h and viral supernatant was harvested after additional 24h. Viral supernatants were filtered through a 0.45 µm filter and applied to target cells for 24-48h. Retroviral particles were produced in HEK-GP cells following the above protocol. Plasmids expressing wildtype or mutant (S710F) STAT5A under control of the murine stem cell virus promoter (MSCV) or empty control were kindly provided by Dr Richard Moriggl. The knockdown of CD271/NGFR was performed with a DOX-inducible shRNA (SMARTvector, Dharmacon, clone ID V3SVHS02_8785341). Transgene expression was induced and maintained with DOX at a final concentration of 4 µg/ml. Virally transduced cells were selected for puromycine (Puro) resistance using a final concentration of 10 µg/ml Puro. Stable selection was achieved after passaging and growth of cells in presence of Puro for ∼3 passages.

### Methylome profiling

For global methylome analysis, DNA of snap frozen or FFPE MBM was isolated according to standard procedures using the DNeasy blood and tissue DNAextraction kit (Qiagen, Hilden, Germany). 500 ng genomic DNA were subjected to bisulfite conversion using the EZ DNA Methylation-Gold Kit (Zymo Research) according to the manufactureŕs protocol. Subsequently, samples were analysed on the Infinium MethylationEPIC Kit (Illumina) according to the manufactureŕs recommendations to obtain genome-wide data from 850,000 CpG positions. Raw data from Illumina Epic arrays were preprocessed and analyzed in the standard workflow of the packages RnBeads^51^ and watermelon^52^. Differential methylation analysis was conducted on site and region level according to the sample groups regarding their levels of E-cadherin or NGFR expression or level of immune cell infiltration (TIL status) or mutation status of BRAF. For statistical analysis, p-values on the site level were computed using the limma method. I.e. hierarchical linear models from the limma package were employed and fitted using an empirical Bayes approach on derived M-values.

### Gene-set enrichment GSEA/Single-sample GSEA

GSEA was performed using the most current BROAD javaGSEA standalone version (http://www.broadinstitute.org/gsea/downloads.jsp) and gene signatures of the molecular signature database MsigDB^53, 54^, 7.4 (Hallmark, C2) as well as published signatures specifying different phenotypic states of melanoma such as “Melanoma aggressiveness^55^”, “Proliferation”, “Invasion”^56^, parts of the IPRES signatures (“MAPKi-induced EMT^57^”) and MITF-target gene signature (“MITF_Targets_TCGA^58^”). The NGFR^high^ gene signature that defines anti-PD-1 therapy resistance was kindly provided by Oscar Krijgsman and Daniel S. Peeper^59^ and overlaps with our identified set of CD271-associated genes^32, 33^. Gene signatures defining the undifferentiated neural-crest cell state were taken from Tsoi et al.^60^ Analyses of single signatures were run using 10,000 permutations; analyses of signature collections were run using 1,000 permutations. Genes were ranked based on the Signal2Noise metric. Ecad- and CD271-associated gene signatures were defined by the comparative analysis of Ecad^high^ or CD271^high^ vs low MBM.

### Confocal microscopy

High-resolution immunofluorescence imaging of tumor sections and cell lines was performed with an LSM880 airyscan confocal microscope (Zeiss) and appropriate software (Zen black, ver. 2.3 SP1). Images were taken with objectives 10x, 20x and 63x/1.40 plan-apochromat, oil dic M27) at a resolution of 2048x2048 pixels/cm, 8bit, scan speed 6, averaging 4. Imersol 518F was used for oil microscopy. Stacked multichannel image files (czi) were separated and background adjusted using AdobePhotoshop2020 and stored as merged tiff files at a resolution of 600 dpi. Z-stacks were converted into three-dimensional images using the arivis tool of ZEN2 software.

### 3D-invasion assays

Briefly, 50 µl of ice cold matrigel (Corning, 734-0270) were plated per well of a cooled 96-well plate and incubated for 10 min in a standard cell culture incubator at 37 °C. After matrigel polymerization, 2,500 cells of BMCs were plated on top of the matrigel layer in 100 µl medium. Images were taken every 3 days for tracking of spheroid formation.

### Live cell imaging-based migration assays

The migratory capacity of unmodified or modified shRNA or reporter expressing BMCs was assessed using the Incucyte® Zoom live-cell imaging system. Briefly, 3x10^4^ cells/well of each cell line were seeded on 96-well plates 24 h before, yielding a dense cell layer. Reproducible scratches were performed using the Incucyte® WoundMaker tool (EssenBioscience/ Sartorius) and floating cells were removed by gentle washing of wells with medium.

After wounding, the 96-well plate was placed into the live-cell imaging system and cell migration was monitored every 4h for 7 days, using a × 10 magnification. Serial pictures were stacked for movie preparation using the ImageJ software (https://imagej.nih.gov/ij/). Statistical analysis was performed by using a two-tailed, paired t-test.

### Western blotting

Whole protein was isolated from frozen cell pellets using RIPA buffer and protein concentration of lysates was determined by Bradford assays (Pierce™ Coomassie Plus Assay Reagent, Thermo). 25–40 µg of total protein lysates were separated on 12% SDS-PAGE gels and transferred to PVDF membranes (Merck) by using the turbo semi-dry blotting system (BioRad). Membranes were blocked with 5% BSA solution and incubated with primary antibodies (CD271, clone D4B3, XP, Cell signaling; and β-Tubulin, clone 9F3 both from Cell Signaling Technology, Germany; all diluted 1:1000) overnight at 4°C. For signal detection membranes were washed twice with PBS-Tween20 (0.1%) and incubated with a horse radish peroxidase (HRP)-coupled secondary antibody (goat anti-rabbit IgG, Cell signaling) for 1 h at RT and analyzed with an automated imaging system (Vilber).

### Animal experiments

All experiments with animals were performed in accordance with the German Animal Protection Law under the permission number G0130/20 obtained via the Berlin Ministry of Health and Social Affairs (LaGeSo). ARRIVE 2.0 Guidelines were strictly followed. Female CD-1 nude mice (8-9 weeks of age, 24-26g, Charles River Laboratories) were stereotactically inoculated with 2.5x10^4^ BMC1-M1 and BMC1-M4 cells using a 1µl Hamilton syringe and a stereotactic frame as described previously^61^. Briefly, the bur hole was placed 2 mm lateral (right) and 1 mm rostral from the bregma. The cells were administered at a depth of 3 mm. The number of cells used for the inoculation was determined in accordance with previous literature with the established human melanoma cell line M14^62^. For the procedure, the animals received anesthesia (9mg Ketamine-Hydrochloride (CP-Pharma Handelsgesellschaft mbH, Burgdorf, Germany) + 1mg Xylazine (CP-Pharma Handelsgesellschaft mbH, Burgdorf, Germany) per 100g) intraperitoneally as well as subcutaneous prophylaxis against infection (10’000 I.E, benzylpenicillin potassium, InfectoPharm Arzneimittel und Consilium GmbH, Heppenheim, Germany) and analgesia (100 mg/kg Paracetamol (B. Braun Deutschland GmbH & Co. KG, Melsungen, Germany), Lidocaine (Aspen Germany GmbH, Munich, Germany). Additionally, analgesia (300 mg/kg*d Paracetamol, bene-Arzneimittel GmbH, Munich, Germany) was administered via the drinking water for the first two postoperative days. Following the procedure, MRI scans were performed every 7 days until either the tumor volume was above 20 mm³ or at latest on the 49th day after implantation. Animals were sacrificed by perfusion with 4% PFA in deep anesthesia. The animals were kept in a 12-hour light-dark cycle and had ad libitum access to water and food. Examinations for general and neurological symptoms were performed once a day and on the first two postoperative days twice a day. The weight was measured postoperatively once a day followed by regular weight monitoring once a week.

### Intracranial injections of A375 cells

For assessment of the proliferative capacity of non-metastatic melanoma cells (A375) 1x10^3^ cells/µl were injected into the right hemisphere of NOD scid gamma (NSG) mice, carried out by EPO GmbH (Berlin, Germany). Tumor growth was tracked by bioluminescent imaging. Mouse brains (n=3) were formaldehyde fixed and embedded after ∼25d. Brain sections (2 µm) were processed for immunofluorescence and confocal imaging as described.

### Magnetic Resonance Imaging (MRI)

The MRI scans were performed using a 7 Tesla small animal MRI (BioSpec 70/20USR or PharmaScan 70/AS, Bruker Biospin, Ettlingen, Germany and ParaVision 6.0.1 or 5.1 software). During the scans, the mice received inhalation anesthesia (1.0-1.5% Isoflurane (CP-Pharma Handelsgesellschaft mbH, Burgdorf, Germany) in a mixture of 30% oxygen and 70% nitrous oxide). The depth of the anesthesia was monitored using the respiratory frequency (70-120 breaths per minute). T1 weighted sequences (TR = 1,000 ms, TE = 10 ms, RARE factor = 2, 3 averages for BioSpec; TR = 975 ms, TE = 11.5 ms, RARE factor = 2, 4 averages for PharmaScan) after intraperitoneal administration of gadolinium-based contrast agent (12,09 mg per mouse in a solution with 180 µl 0,9% NaCl, Gadovist, Bayer AG, Leverkusen) and T2 (TR = 4,200 ms, TE = 36 ms, RARE factor = 8, 3 averages for Biospec and TR = 4,200 ms, TE = 36 ms, RARE factor = 8, 4 averages for PharmaScan) were measured. The tumor volume was measured using ITK-SNAP 3.8.0 Software^63^ (Paul A. Yushevich, Guido Gerig, www.itksnap.org).

### Quantitative real-time PCR

RNA isolation from frozen cell pellets was performed with the RNeasy Mini Kit (Qiagen, Germany) and, following the manufacturers protocol. Reverse transcription of 500 ng–2.5 µg RNA was performed with SuperScript VILO cDNA synthesis kit (Invitrogen, Germany) and diluted to a final volume of 50 µl. qRT-PCR was carried out on a Step one plus PCR cycler (Applied Biosystems, Germany) for 30–40 cycles. Primers were designed for 55–60°C annealing temperatures. Relative expression levels were calculated with the CT method^64^, normalized to β-actin. Primer sequences are shown in Supplementary Table 9.

### Drug sensitivity assays

The response of BMCs and conventional melanoma cell lines to dabrafenib, tivantinib in a range of 1nM-10µM and paclitaxel in a range of 1pM-10nM of eight technical replicates was determined densitometrically after 72 h and fixation/staining with crystal violet. Drug treatments were performed 24 h after seeding of 5,000 cells/96-well. Live-cell imaging-based drug responses were performed accordingly. Images were taken every three hours using a 10x objective and the general label-free mode, two pictures of eight technical replicates per condition were taken. Drug response was assessed by changes in the cellular density over time. The cell density was determined by a confluence mask tool as part of the Incucyte software. IC50 values were calculated by curve-fitting (https://search.r-project.org/CRAN/ refmans/REAT/html/curvefit.html) based on confluence measurements at day 3.

### Data deposit

Whole transcriptome and methylome data were deposited in the European Genome-Phenome Archive (EGA), under accession numbers EGAS00001005976 and EGAS00001005975 (https://ega-archive.org/).

## Results

### Therapeutic interventions promote the development of invasive brain metastases

The investigation of longitudinal and synchronous BM can provide insights into the processes that control clonal and subclonal evolution, the driving forces of tumor progression. Metastatic melanoma is still a devastating disease as patients with stage IV melanoma present with metastases at multiple extracranial and intracranial sites^65, 66^. Presumably, the development of extracranial metastases precedes BM formation (Fig. 1a). Several lines of evidence suggest that therapeutic interventions enhance the emergence of therapy-resistant cellular subclones driving relapse and intracranial progression of MBM^67–69^. To ascertain whether therapeutic interventions promote the development of migratory and invasive tumor cell phenotypes, we investigated the levels of CD271/NGFR of extensively treated (BRAFi/MEKi and radiation or radiation and ICi) but progressive MBM. We observed high expression of CD271 (70–100%) throughout the entire tumors (Fig. 1b, left panels and Supplementary Fig. 1a) irrespective of therapeutic interventions, mutation status (BRAF, NRAS) and intracranial location of MBM but potentially promoted by reactive astrocyte secreted factors. The BRAF/NRAS mutation status of tumors was determined by TargetSeq during routine diagnostic work-up. Whole transcriptome data of MBM prior and after BRAFi/MEKi therapy are not available. Therefore, we investigated the levels of CD271 in a set of drug-naïve, prior- (pre-relapse) treatment melanoma and tumors that relapsed after dabrafenib/trametinib (GSE77940^70^) treatment (post-relapse), (Fig. 1b, right panel). CD271/NGFR was significantly (2.5fold, p=0.034) increased in four of five post-treatment tumors. Gene-set enrichment analysis (GSEA) revealed an invasive phenotype of post-treatment tumors and a higher representation of signature genes indicating an undifferentiated neural crest (NC)/NCSC-like (NES=2.953, FDR<0.001) or CD271/NGFR- driven (NES=2.646, FDR<0.001) or invasive (NES=2.284, FDR<0.001; Hoek signature^56^) tumor cell state (Fig. 1c). Concordantly, we observed decreased levels of E-cadherin (CDH1, -0.212fold, p=0.036), (Supplementary Fig. 1b). Relapsing tumor cells featured enrichment of previously defined CD271-responsive genes (NES=2.646, p<0.001) and a MAPKi-induced EMT-like (NES=1.911, FDR<0.001) phenotype (Supplementary Fig. 1c). As we hypothesized that CD271-driven programs mediate the progression of MBM by fostering the emergence of micrometastases, we investigated matched pre- and post-relapse tumors. We observed that CD271^+^ cells infiltrated the brain tumor environment, BTE (Fig. 1d, left panels) and formed micrometastases in close proximity to MBM/BTE transition sites (Fig. 1d, right panels).

**Fig. 1:**
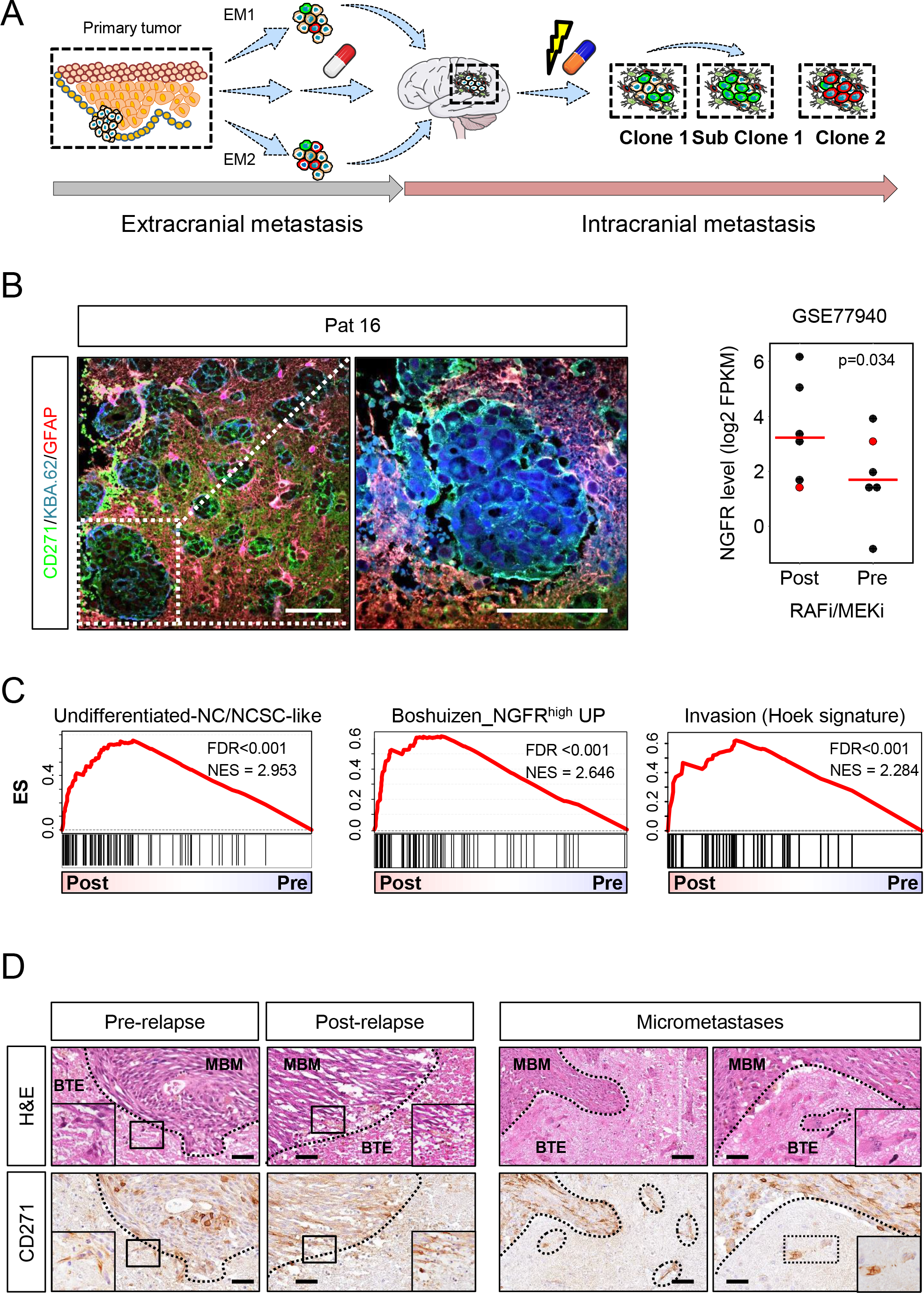
MAPKi treatment increases invasiveness of melanoma. a.) Schematic representation of the metastatic cascade establishing extracranial (EM1, EM2, EMn, etc.) and intracranial metastases and routes of intracranial metastasis leading to sub clonal evolution. Therapeutic interventions likely foster extracranial metastasis and intracranial progression by subclonal evolution. b.) Immunohistochemistry (IHC) of CD271 in a representative MBM of patients who received combinatory therapies: BRAFi/ MEKi (dabrafenib/ trametinib), ICi (nivolumab and/or ipilimumab) or radiation therapy. Scale bars indicate 50 µm. Right panel: NGFR expression levels of melanoma pre- and post RAFi/MEKi therapy (study GSE77940), one tumor has been excluded (red filled circle) in each group. T-test was used to calculate significance. c.) GSEA of pre- and post-treatment melanoma revealed enrichment of undifferentiated, neural crest (NC)-like, anti-PD-1 resistant/ NGFR^high^ and invasive cells in post-treatment melanoma. False-discovery rate (FDR) indicates significance of analysis. d.) IHC of matched initial and relapsed tumors revealed CD271^+^ tumor cells that showing infiltration of the brain tumor environment (BTE) and formation and micrometastases. Hematoxylin and eosin (H&E) staining shows discrimination of tumor (MBM) and BTE. Scale bars indicate 50 µm.

Mechanisms of early brain development are conserved among vertebrates. However, recent single-cell sequencing studies of the developing human brain revealed spatial differences of the cellular composition^71^. Hence, the response of tumors to environmental cues is likely determined by the cellular composition of environmental cells and secreted soluble factors, programing progression stages of MBM and primary brain tumors^72^. To identify molecular subsets that potentially reveal the progressive state of MBM, we collected intraoperative and cryo-preserved MBM (n=16; Supplementary Table 1) from different intracranial sites (Fig. 2a) including longitudinal metastases (Pat8) and patient matched pairs (patients 23, 24). The latter synchronous metastases of patient 23 that developed within the occipital and parietal lobes or MBM of patient 24 that emerged within the frontal lobe and lobus insularis, like metastases of Pat8 (M2-M4) featured a therapy-resistant phenotype (Fig. 2a). Whole transcriptome profiling of MBM and normal brain controls (Cortex, Pons, Cerebellum/Cereb; BC; Supplementary Table 2) rather revealed a separation into subgroups regarding the content of admixed brain parenchyma and irrespective of the intracranial region of tumors or genetic state (presence of BRAF or NRAS mutations) (Fig. 2b). As resection of MBM usually includes a margin of adjacent parenchymal cells, we determined the differentially regulated genes (DEGs) among BC and MBM and identified a pan-gene signature potentially defining MBM showing enrichment of extracellular exosomes and genes that are associated with focal adhesion or defining extracellular matrix components (ECM) or melanosomes (Supplementary Fig. 2a). Classifiers suggest a different distribution of molecular subsets among the main subgroups such as the enrichment of classifiers of an NGFR^high^ or invasive subset. Moreover, we observed a strong homogeneity of matched, synchronously resected metastases of patient 23 and a clear separation of metachronous (2018, 2020), drug-responsive or drug-resistant subclones M1 or M4 of Pat 8. The initial TargetSeq of hot spot regions of a panel of 50 cancer-associated genes provided information about the BRAF/NRAS status of MBM (n=37) and identified genetic aberrations in 11 genes among them expected drivers of melanoma progression such as CDKN2A (Supplementary Fig. 2b- c). Generally, we discovered that tumors were either BRAF or NRAS mutated.

**Fig 2:**
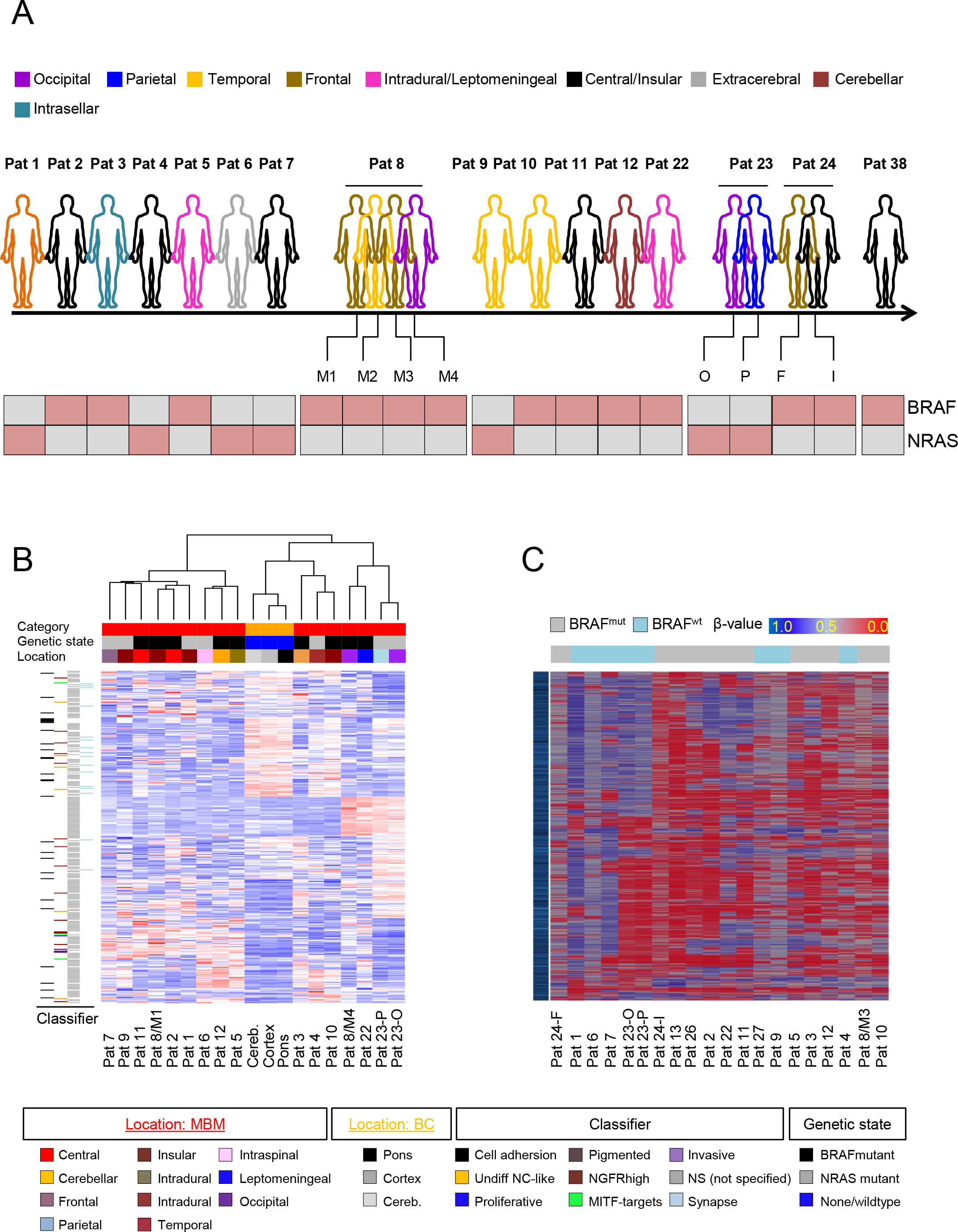
Characterization of MBM revealed different molecular subtypes. a.) Schematic overview about melanoma patient-derived MBM samples regarding their concordance, intracranial site and BRAF/NRAS mutation status. b.) Supervised clustering of MBM (n=16) and BC (n=3) revealed clustering independent from intracranial sites of discordant MBM or genetic state. Classifiers indicate the presence of tumors featuring an undifferentiated-neural crest (NC)-like or pigmented, invasive, proliferative, NGFR^high^ state or expressed MITF- targets or genes involved in cell adhesion represented in the top1000 variably expressed genes. c.) Heat map representing the top1000 variably methylated CpG islands among MBM, BRAF mutations status is indicated.

Global methylome profiling serves as a tool for the comprehensive molecular classification of several primary brain tumors^73, 74^. We used 850k methylome profiling to gain insights into the epigenetic landscapes of MBM (n=20) and to assess potential molecular subgroups. Moreover, we asked for the similarity of concordant tumors that were synchronously resected from different regions within the brain. The comparison of synchronous matched MBM revealed a clear simultaneous clustering of occipital and parietal tumors of patient 23 (Fig. 2c), in concordance with whole transcriptome data. On the other hand, we observed a divergent clustering of frontal and insular tumors of patient 24. Therefore, even though matched tumors likely have synchronously evolved from a common founder clone during the course of treatment and were simultaneously resected, MBM might follow very different developmental routes. To exclude that the clustering was affected by admixed brain-derived stromal cells, we calculated the tumor cell content based on the expression levels of PRAME (Preferentially Expressed Antigen in Melanoma) observed in melanoma cell lines. PRAME was coherently expressed in MBM (median: 79.8%, range: 119.4-72.8%) with one exception (Pat 10) that showed no PRAME expression and was classified as primary central nervous system (CNS) melanoma. PRAME was not expressed in brain-derived stromal cells and several MBM exhibited even higher levels in comparison to melanoma cell lines (Supplementary Fig. 3a, left panel).

Next, we wondered whether the subsets Ecad^high^ vs low, TIL^high^ vs low and BRAF^mut^ vs wt may be further classified by a set of differentially methylated genes (DMGs). We did indeed find a significant (FDR-adjusted p-value<0.05) difference of 46 CpG islands in promoters of 35 genes that were hypomethylated in BRAF^mut^ tumors (Supplementary Fig. 3a, right panel). The matching with transcriptome data revealed 14 MBM expressed genes that featured high methylation of promoters in BRAF^wt^ tumors and identified integrin b7 (ITGB7) as a potential predictor of favorable survival (Supplementary Fig. 3b-c).

### The progression of MBM accompanies an E-cadherin-to-CD271 phenotype switch

Non-genetic processes such as cellular plasticity and phenotype switching are driving forces of tumor heterogeneity and likely determine drug response and tumor relapse^75^. Several lines of evidence suggest that therapeutic interventions blocking oncogenic BRAF (BRAFi/MEKi) and/or inflammation-induced processes subsequently activate mechanisms driving invasive and migratory properties of melanoma cells. These are associated with tumor relapse and critical for the emergence of MRD^3, 23, 24, 28, 35, 76, 77^. To gain insights into phenotypical and molecular changes that occurred alongside progression, we investigated spatially separated, longitudinal metastases of Pat8 that were collected before BRAFi/MEKi therapy (M1) or which have developed and progressed under therapy (M2, M3, M4) (Fig. 3a). We observed a high level of CD271 expression in M3 and M4 but a low level in pigmented subclones M1 and M2 (Fig. 3b, left and center panels and Supplementary Fig. 4a). At the time of M4 resection, the patient exhibited a very aggressive disease stage that was accompanied by *meningeosis melanomatosa,* the penetration of (HMB45 positive) melanoma cells into the CSF (Fig. 3c), suggesting that the emergence of CD271-driven subclones likely indicated a progressive intracranial disease stage.

**Fig 3:**
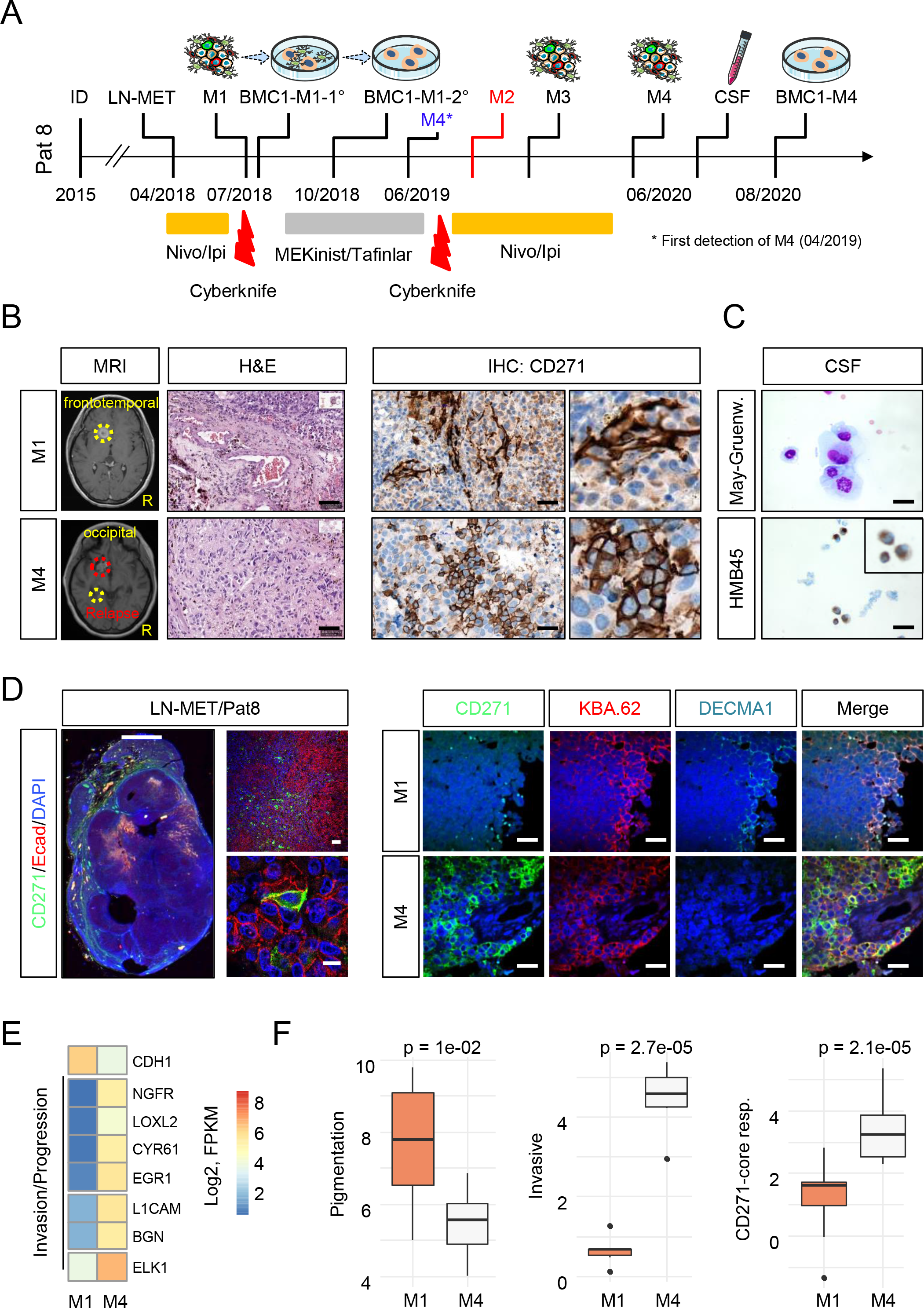
Intracranial progression accompanied an Ecad^+^ to CD271^+^ phenotype switch. a.) Timeline indicating the therapeutic history and time points of surgical removal of tumors, cerebrospinal fluid (CSF) and establishment of tumor-derived cell lines of Pat8, initially diagnosed with primary melanoma in 2017. b.) Magnetic resonance imaging (MRI) of M1 and M4, reflecting disease progression (left panels). Hematoxylin/eosin (H&E) and immunohistochemistry (IHC) for CD271 indicates rare CD271^+^ cells in M1 but a high level of CD271 expression in M4 (center and right panels). c.) May-Gruenwald (May-Gruenw.) staining and IHC for HMB45 indicates the presence of MBM cells in CSF (right panels). d.) IF of a M1 concordant lymph node metastasis (LN-MET) for Ecad and CD271/p75^NTR^, indicating unique and co-expression of both (left panels). Comparative imaging of M1 and M4 for levels of cell surface expression of Ecad/ DECMA-1 and total levels of CD271. KBA.62 served as marker of stem-like and non-stem like human melanoma cells (right panels). Scales in (b-d) indicate 50 µm. e.) Expression levels (Log2FPKM) of Ecad (CDH1), NGFR and additional markers of invasion and progression, illustrate the difference of phenotypes of M1 and M4. Levels are color coded. f.) Box plots indicate the levels (mean±sdv) of genes indicating pigmented, invasive of CD271/NGFR-driven tumor cells in M1 and M4. T-test was used for calculation of p-values.

A comparative analysis of whole transcriptomes of M1 and M4 revealed a high concordance (R=0.85; p<2.2e-16) and uncovered ∼1,000 differentially regulated genes probably indicating different evolutionary traits of subclones (Supplementary Fig. 4b, Supplementary Table 3). Our survey identified decreased levels of E-cadherin (Ecad, ∼5fold) and of pigmentation markers (DCT, TYR, MLANA ∼7fold) and showed CD271/NGFR among the top upregulated genes in M4 (∼25fold). Presumably, the Ecad-to-CD271 phenotype switch marks a final step of MBM progression, which is rather a stepwise and slowly proceeding than a rapid process. We examined the levels of Ecad and CD271 in a lymph node metastasis (LN-MET) and concordant MBM (M1, M4) and validated co-occurrence of melanoma cells that featured distinct (∼35% CD271^+^; ∼50% Ecad^+^) and overlapping (∼10% CD271^+^/Ecad^+^) expression of Ecad and CD271 in LN-MET (Fig. 3d, left panels and Supplementary Fig. 4c). Moreover, we observed distinct expression of both markers in M1 and M4 (Fig. 3d, right panels). CD271^+^ cells were rarely found in Ecad^+^ positive M1 (Supplementary Fig. 4d-e) and CD271^+^/Ecad^+^ cells that potentially reflected the plasticity-driven transition of Ecad^+^ into CD271^+^ cells were not evident in M4. KBA.62 was used as a general marker of melanoma cells that enabled detection of stem-like CD271^+^ and non-stem-like melanoma cells of primary and metastatic melanoma^78^. Next, we sought for additional markers of invasive and progressive tumor cell phenotypes such as LOXL2 and CYR61 among the set of M1/M4 differentially regulated genes and observed a clear separation of both tumors regarding the gene expression levels (Fig. 3e). Moreover, we applied gene signatures characteristic of CD271-dependent or invasive programs and observed a significant higher median expression of CD271-dependent (p=2.7e-05) and invasion-associated genes (p=2.1e-05) and a reduced expression of pigmentation-associated markers (p=1e-02) in M4 vs M1 (Fig. 3f). Ecad expression in melanoma was associated with a proliferative and pigmented cell phenotype. Thus, our findings strengthen the concept of a proliferative-to-invasive phenotype switch that likely promotes MBM progression.

### E-cadherin and CD271 define molecular subgroups of MBM

Ecad mediates the junctional connection of melanocytes and keratinocytes. The malignant transformation of melanocytes to melanoma accompanies the downregulation of Ecad that imply a low level of Ecad expression in metastases. We identified Ecad among the top 1000 variably expressed genes in MBM and by the direct comparison of BC vs MBM (FClog2=8.2, padj=0.012, Supplementary Table 2). Next, we analyzed for the frequency of Ecad- and CD271-expressing MBM and observed that 56.3% (9/16) of MBM were Ecad positive. Levels of Ecad expression of another patient with *meningeosis melanomatosa* (Pat 11) and concordant pairs of pre- vs. post-relapse (Pat 19) and extracranial (spinal) vs. intracranial (Pat 6) MBM (Fig. 4a) were comparable. The latter finding was confirmed by a comparison of whole transcriptome data (EGAS00001003672^79^) of intracranial (n= 79, BM) and extracranial metastases (n=59, EM; p=0.6144), (Supplementary Fig. 5a, left panel). Likely, brain metastatic tumor cells exhibit a rapid EMT-MET capacity and metastases re-acquire Ecad expression soon after every metastatic step. We surveyed the TCGA data set comprising primary melanoma (PT), EM and a rare subset of BM (n=6) and observed lowest levels of Ecad in EM (Supplementary Fig. 5a, right panel).

**Fig. 4.**
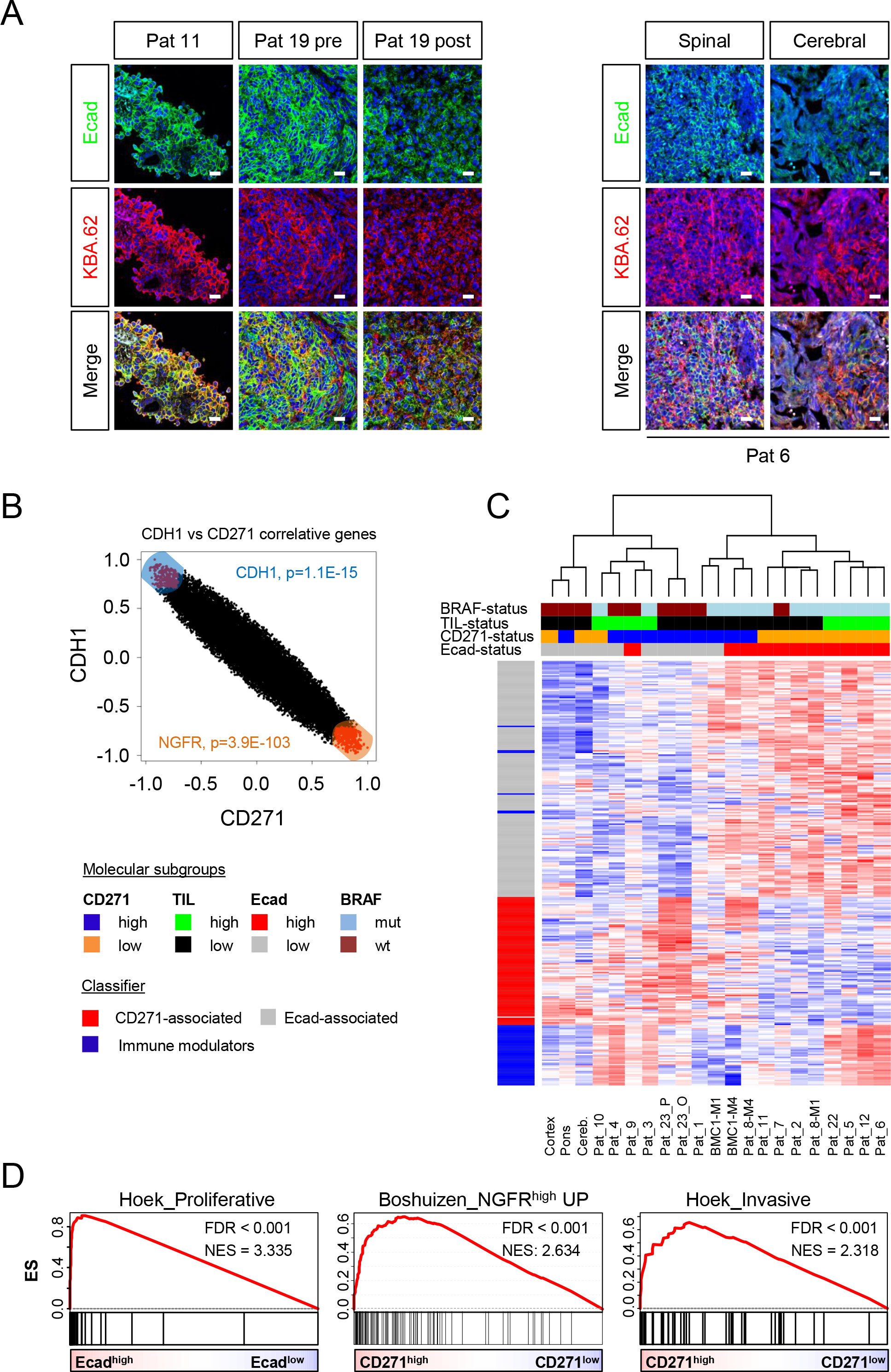
Expression of E-cadherin and NGFR defines molecular subgroups of MBM. a.) IF and confocal microscopy of epithelial-like MBM (Pat 11) and a matched pair of initial (Pat 19, frontal lobe, left) and relapsed (Pat 19-R, cerebellum, left) tumors for Ecad and KBA.62 revealed intense staining and proper membrane localization of Ecad (∼80% of cells) in Pat 19 but a reduced level (∼50%) and punctuated localization in Pat 19-R (left panels). IF of spinal and concordant cerebral metastases demonstrated the maintenance of Ecad expression in extra- and intracerebral metastases. Bars indicate 50 µm. b.) Association of Ecad/CDH1 and CD271/NGFR correlated genes displayed a mutually exclusive pattern. c.) Molecular subclustering of MBM regarding the expression of Ecad- and CD271-associated or immune modulatory genes. Additional molecular subgroups (BRAF^mut^ vs. wt; TIL^high^ vs. low) and levels of Ecad and CD271 expression are color coded. Heat map presents a supervised, euclidean and ward.D clustering. d.) GSEA of Ecad^high^ and CD271/NGFR^high^ subsets for representation of gene signatures specifying a proliferative, NGFR-driven or invasive phenotype of MBM. FDR indicates the significance of enrichment; ES, enrichment score; NES, normalized enrichment score. 10,000 permutations were performed.

We ranked MBM regarding Ecad/CDH1 and NGFR levels (Supplementary Fig. 5b-d) and correlated genes, that are associated with either Ecad (1,803 genes, p ≤ 0.05) or NGFR ≤ 0.05) expression (Supplementary Table 4) and found an inverse association of Ecad and NGFR correlated genes (Fig. 4b). The determination of DEGs among Ecad^high^ and Ecad^low^ subgroups revealed CD271/NGFR and CRABP2 among the top expressed genes in the Ecad^low^ (NGFR^high)^ tumors and SLC45A2, TSPAN10 and ABCC2 expressed by Ecad^high^ tumors (Supplementary Table 4). Moreover, we validated that MITF-targets were positively or negatively correlated with Ecad or NGFR/CD271 tumor cell subsets (Supplementary Fig. 5e). Next, we determined the association of both molecular subsets Ecad^+^ and CD271^+^ with BRAF mutation status. We validated the representation of gene signatures in MBM and observed that nearly all MBM with IHC-proven Ecad^+^ or CD271^+^ phenotypes featured expression of signature genes (Fig. 4c). We discovered a correlation of CD271/ Ecad expression, with BRAF status, suggesting that BRAF^wt^/NRAS^mut^ MBM feature increased levels of NGFR/CD271 (p=0.014) and BRAF^mut^ MBM are accompanied by Ecad expression (p=0.013, Supplementary Fig. 6a). In addition, single sample GSEA (ssGSEA) revealed a correlation of CD271^high^ MBM with brain metastasis in breast cancer (p<0.05, Supplementary Fig. 6b-c, first panel) and validated the NGFR/p75^NTR^ dependency of CD271^high^ tumors.

Besides Ecad and NGFR expression, the presence of tumor-infiltrated lymphocytes (TILs) served as a subset classifier of MBM (Fig. 4c). Generally, primary and secondary brain tumors are immunologically cold (non-inflamed) tumors^80^ and efficiently evade immune surveillance. Though a subset of MBM in our cohort (n=8; 50%) showed infiltration of CD3^+^ T cells even discriminating M1 and M4 (Supplementary Fig. 6c, center, right panels) and clearly distinguished favorable^81^ TIL^high^ and TIL^low^ subsets by expression levels of CD3D (p=2.8e-07) and CD8A (p=2.6e-04; Supplementary Fig. 6d). Moreover, TIL^high^ tumors featured increased inflammatory responses and activation of astrocytes (Supplementary Fig. 6e), suggesting that inflammatory processes involving emergence of reactive astrocytes precede immune cell infiltration of tumors^82^. GSEA unraveled the molecular features of Ecad^+^, CD271^+^ or TIL^high^ tumors (Supplementary Table 5) and revealed a proliferative phenotype (Hoek_Proliferative, NES = 3.335, FDR<0.001) of Ecad^pos^ tumor cells. Ecad^low^/CD271^high^ tumors featured enrichment of a tumor-intrinsic NGFR signature that was derived from a set of melanoma cell lines which had spontaneously acquired resistance to T cells (Boshuizen_NGFR^high^_UP, NES=2.634, FDR<0.001). The signature potentially predicts anti-PD-1 therapy resistance, and increased immune exclusion. Moreover, Ecad^low^/CD271^high^ MBM exhibited an invasive phenotype (Hoek_Invasive, NES = 2.318, FDR<0.001) among other core enrichments (Fig. 4d and Supplementary Table 5).

Hence, although the emergence of MBM is generally associated with poor prognosis, the different phenotypes might determine the degree of aggressiveness of intracerebral tumors and their capability of formation of multiple brain metastases and response to therapeutic interventions.

### MBM-derived cell lines reflected and maintained tumor-cell properties

The mutual interaction of MBM cells with components of the brain microenvironment such as growth factors, chemokines and extracellular matrix (ECM) proteins probably has a strong impact on disease progression. Very recently, the release of CD271-carrying extracellular vesicles was recognized as a critical step in lymphangiogenesis and metastasis^37^. Moreover, neurotrophins such as NGF and BDNF that are released by reactive astrocytes likely induce a migratory and invasive phenotype of MBM^83^. Besides the supportive function of the tumor micronenvironment for the establishment and progression of MBM, we wondered whether the progressive state of MBM cells is maintained by *in vitro* conditions. Therefore, we established a panel of stable cell lines from intraoperative MBM of patients that had not (BMC1-M1, BMC2) or had received (BMC1-M4, BMC4) BRAFi/MEKi or ICi therapies. Intraoperative MBM from different brain regions were minced and enzymatically digested to obtain a single cell suspension that predominantly contained tumor cells but also admixed environmental cells such as astrocytes, microglia and oligodendrocytes (Fig. 5a, 1°). The passaging of 1° cells subsequently enriched for tumor cells that adapted to the *in vitro* conditions. All patients exhibited multiple BM at the time of operation; hence, we expected MBM-derived tumor cells to feature a high migratory and invasive phenotype, which generally have been associated with the expression of markers such as AXL, MET and CD271. However, only minor cellular subsets featured AXL (2.1%/1.6%/0.5%) and MET (1.7%/36.7%/0.4%) expression in BMC1-M1, BMC1-M4 and BMC4 cells. Nonetheless, CD271 was highly expressed in adherently growing BMCs and associated suspension cells (Figs. 5b; 5c, right panels and Supplementary Fig. 7a-b). The majority (3/5; 60%) of BMCs featured a suspension phenotype that was likely related to a high CD271 level^84^ and associated mutations. High levels of AXL had been attributed conferring a migratory and invasive phenotype to melanoma^85^. We detected a significantly higher AXL expression in MBM than BC (Supplementary Fig. 7c), validated in a subset of AXL^+^ MBM and BMC1-M1 cells (Supplementary Fig. 7d). However, we found a discordant pattern of expression of AXL and its ligand GAS6 in MBM (not shown) and rarely observed activation/ phosphorylation of AXL (Supplementary Fig. 7e).

**Fig. 5:**
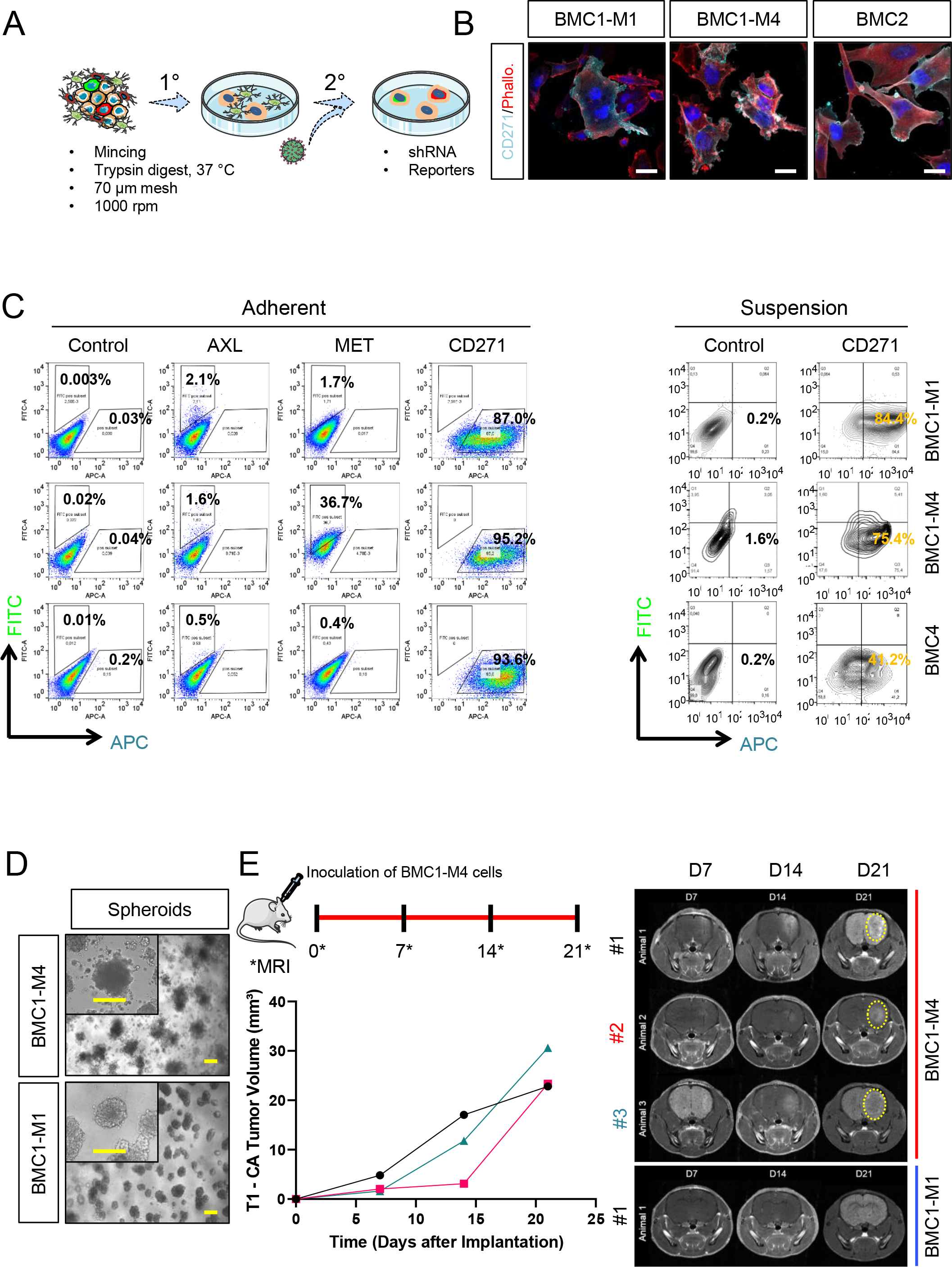
MBM-derived cell lines serve as *in vitro* models and exhibited distinct migratory phenotypes. a.) Simplified experimental scheme of MBM-establishment, 1° tumor cell cultures contained admixed cells such as astrocytes, which were subsequently lost. 2° cells comprised a heterogeneous mixture of tumor cells that were accessible for genetic modification such as shRNA-based knockdown or generation of reporters. b.) IF of a representative set of BMCs for CD271 and phalloidin (Phallo.), bars indicate 50 µm. c.) Flow cytometric analysis of viable, adherently grown BMCs for cell surface expression of potential drivers of migration AXL, MET (FITC-labeled) and CD271 (APC-labeled, left panels) or cell surface levels of CD271 of suspension cells (right panels). 50,000 cells were monitored. d.) Embedding of BMCs in matrigel established three-dimensional growing spheroids and demonstrated differences of invasive phenotypes. Bars indicate 50 µm. e.) Experimental set- up of *in vivo* experiments (upper panel). CD-1 nude mice (n=3) were inoculated with patient derived cell lines BMC1-M1 or BMC1-M4 on day 0 and the tumor volume of each animal was ascertained over time for up to 21d by MRI in T1 (lower left panel). Representative MRI images taken 7, 14 and 21 days (D) after transplantation depict tumor formation (dotted yellow line) in BMC1-M4 but not BMC1-M1 inoculated and contrasted animals (right panels). The experiments were terminated once a tumor volume of at least 20 mm³ was reached or at latest on day 49.

In response to BRAFi/MEKi, melanoma cells perform phenotype switching, a non-genetic process during which cells convert from a proliferative to an invasive and drug-resistant state^86, 87^. Our previous data suggested that the proliferative and pigmented tumor cell state was represented by Ecad expression, while the migratory and invasive phenotype was characterized by expression of CD271/NGFR. Accordingly, Ecad/CD271 double positive cells seem to indicate tumor cells in the transitory state. We discovered a proliferative phenotype of the progressive tumor M4 (61.0±11.0% Ki67^+^ cells) and a lower and highly variable level of proliferative cells in M1 (17.1±12.3% Ki67^+^ cells, Supplementary Fig. 8a, left panel). This finding was also reflected by Ki67 levels (BMC1-M1, 48.1±7.0%; BMC1-M4, 52.4±19.4%), and BrdU incorporation (BMC1-M1, 35.0±9.9%; BMC1-M4, 45.5±8.5%; BMC4, 25.5±6.2%) of the corresponding BMCs (Supplementary Fig. 8a, right panel). Next, we assessed the migratory capacity of BMCs by a live-cell imaging-based scratch-wound assay. We observed a high migratory capacity as indicted by a rapid wound closure by BMC1-M4 (669.2±42.8 – 2.4±4.1 µm), BMC2 (515.9±52.0 – 11.4±10.5 µm) and BMC4 (567.4±70.5 –5.8±10.0 µm) but a lower capacity of BMC1-M1 cells (709.2±9.3 – 168.1.4±41.1 µm), p(BMC1-M1 vs BMC1-M4)= 0.004 (Supplementary Fig. 8b-c). The capability of spheroid formation of melanoma cells is associated with stemness that in turn is connected with a migratory and invasive phenotype^88, 89^. To ascertain the spheroid-forming capacity of BMCs, we seeded 2.5x10^3^ BMC1-M1 and BMC1-M4 cells onto a layer of matrigel, which enabled spheroid formation following invasion of the matrigel layer. BMC1-M4 cells formed loosely connected three-dimensional (3D) colonies and satellite colonies (Fig. 5d, upper panels) whereas BMC1-M1 established colonies featured a non-scattered phenotype (Fig. 5d, lower panels). Although this assay reflected an interaction of tumor cells with extracellular matrix forming collagens and laminins, tumor cells might feature different phenotypes in response to a brain-associated microenvironment that is build up by astrocytes and microglia among others. The stronger migratory capacity and 3D satellite-growth pattern of BMC1-M4 cells suggest a higher progressive phenotype compared to BMC1-M1 cells. To assess whether “tumor naïve”, normal brain cells were capable of repressing the progressive state of BMC1- M4 cells, we injected 2.5x10^4^ cells of both cell lines into the right hemispheres of CD-1 nude mice (n=3 per group) and weakly tracked tumor formation via MRI for a period of 49 days (Fig. 5e). BMC1-M4 cells established detectable tumors (median volume of 11.77 mm³ (n=3; range: 3-17 mm³) 14 days post injection (range 7-14d) that featured a constant growth and reached a median volume of 23.35 mm³ (range 22-30 mm³) after 21d. In contrast, BMC1-M1 cells indeed featured a less progressive phenotype and established small size tumors (maximum volume: 2.705 mm³; n=2) 35d after injection (Supplementary Fig. 8d-e). Therefore, the progressive phenotype of BMC1-M4 cells and distinct growth properties of both cell lines were maintained *in vivo* and potentially controlled by intrinsic cues.

### Genetic profiling of longitudinal MBM and BMCs revealed a high genetic concordance and validated subclonal stability

Non-genetic molecular programs such as those controlling the MITF/AXL proliferative/invasive switch, drive tumor heterogeneity and establish MRD, a crucial driver of metastatic progression^35, 90^. MRD and high migratory phenotypes of melanoma cells were correlated with the expression of CD271 and observed in BMCs and in a subset but not all tumors of our MBM cohort. As BMCs were expected to serve as *in vitro* models that reflect tumor cell phenotypes, we asked for the comparability of MBM and BMCs. The comparison of whole transcriptomes of MBM and low-passage (p1-p5) MBM-derived cell lines revealed a high concordance of M1 and BMC1-M1 (R=0.89, p<2.2e-16) and M4 and BMC1-M4 (R=0.90, p<2.2e-16), (Fig. 6a). Moreover, total levels of CD271 reflected the migratory capacity of all BMCs except from BMC4 (Supplementary Fig. 9a). As extracranial and intracranial metastases show comparable molecular characteristics^91^, we further investigated whether BMCs were molecularly distinct from conventional cell lines (A375, MeWo) or a patient-derived cell line that was established from a LN-MET (T2002). In principle, the intracranial injection of 1x10^3^ A375 cells into NSG mice gave rise to tumors within 25d (Supplementary Fig. 9b), hence this cell line was capable of adaptation to the foreign microenvironment and undergoing an *in vitro* to *in vivo* switch. A correlation of MBM, BMCs, BCs, conventional cell lines (A375, MeWo), T2002 and melanocytes based on whole transcriptome data revealed that BMCs clustered with the concordant tumors but not with A375, MeWo or T2002 cells (Supplementary Fig. 9c). Hence, BMCs maintained molecular programs that were distinct from conventional cell lines.

**Fig. 6:**
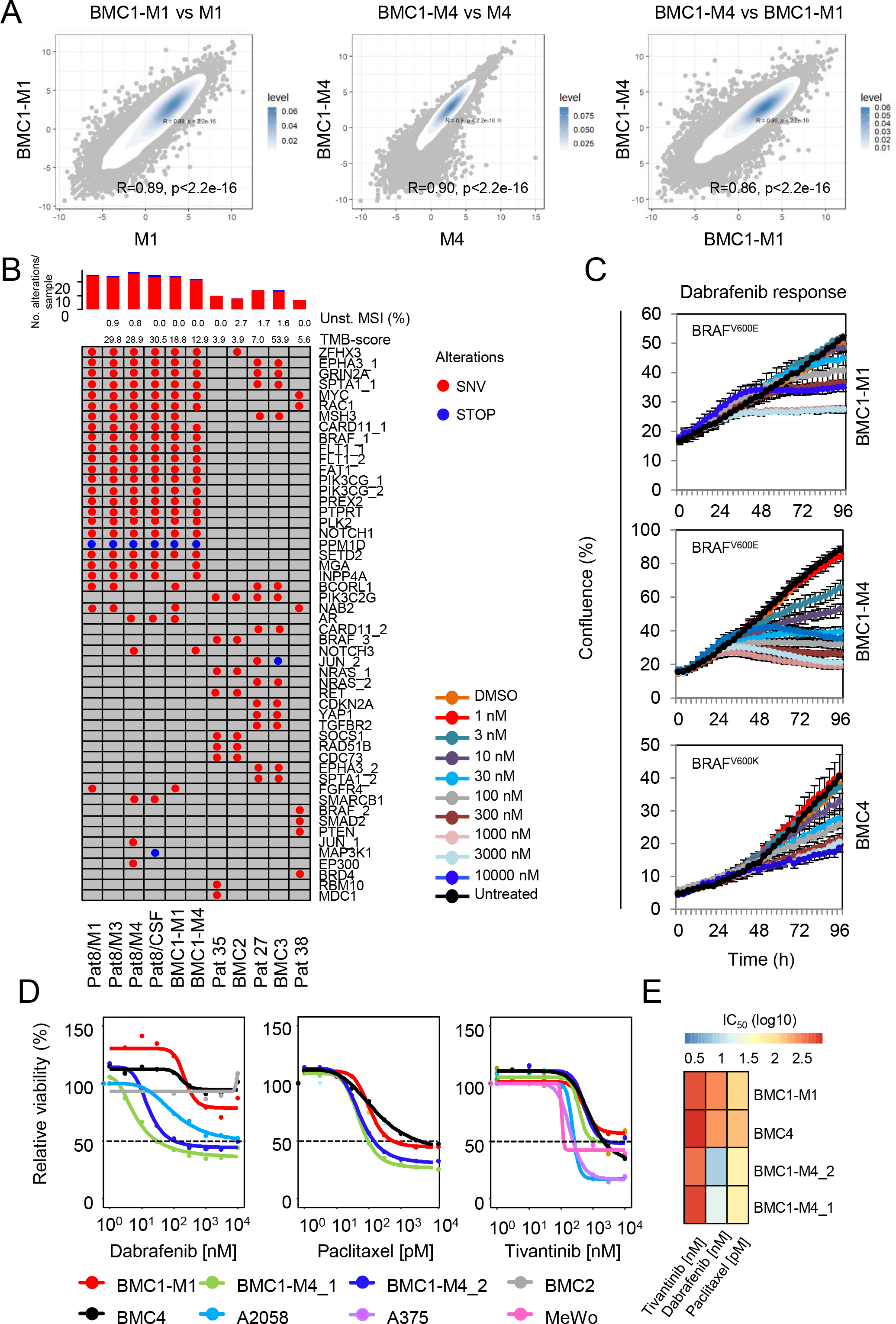
TargetSeq of BMCs and concordant MBM provided insight into the persistence of genetic subclones and subclonal evolution. a.) Scatter plots showing the comparison of BMCs and concordant MBM (∼13,000 genes/sample) indicate a high concordance of transcriptomes of BMC1-M1 vs. M1 (R=0.89, p<2.2e-16) and BMC1-M4 vs. M4 (R=0.90, p<2.2e-16) and narrow differences of BMCs (R=0.86, p<2.2e-16). All p-values were calculated by a two-tailed paired t-test. Levels are color coded and depict levels of concordance. b.) Oncoprint map depicts genetic modifications of 42 genes as determined by high-depth (>775x mean coverage) TargetSeq. Shown are missense (SNVs) and nonsense- mutations (STOP), percentage of microsatellite instability (MSI), level of tumor-mutation burden (TMB-score) and numbers of alterations per sample and per gene. c.) Assessment of sensitivity of BMCs towards increasing doses (1 nM-10 µM) of dabrafenib as determined by live-cell imaging based proliferation indicated by percentage of confluence. d.) Computation of IC50 values by fit-curves of responses of BMCs and conventional cell lines A2058, A375 and MeWo towards dabrafenib (BRAFi), paclitaxel (cytoskeleton targeting drug) and tivantinib (non-ATP competitive inhibitor of c-MET). e.) Depiction of IC50 values of investigated BMCs.

To exclude that MBM-derived cell lines underwent genetic changes triggered by the adaptation to *in vitro* conditions, we performed TargetSeq of longitudinal MBM, BMCs of patient’s 8, 35 and 27 and CSF of Pat8 with a mean coverage of 760x (range 290x-1,505x). TargetSeq provided insights into the permanence of hotspot regions of 560 cancer-related genes and enabled tracing of genetic subclones. The latter potentially emerged during the course of disease in response to therapeutic interventions and microenvironmental changes. We identified 18 ground-state mutations that were commonly found in all specimens of Pat8, particularly BRAF^V600E^ and RAC1^P29S^, which are known genetic drivers of cancer progression^92–95^. Mutations in RAC1 present early UV-radiation caused aberrations, potentially driving BRAFi-resistance, membrane ruffling (Supplementary Fig. 9d) and cell migration^94, 96^. In addition, we identified likely deleterious but functionally uncharacterized mutations in CARD11 and MYC (CARD11^D56N^; COSV62717671 and MYC^N26S^; COSV52371145) that have been associated with cancer^97^ (Fig. 6b and Supplementary Fig. 10a). Apart from public mutations, we detected missense and nonsense mutations, which were exclusively found in either of the longitudinal tumors (Fig. 6b and Supplementary Fig. 10a). These private mutations were persistent; hence, subclones carrying a certain mutation such as FGFR4^S431S^ were maintained *in vitro*. In addition, we observed that subclones bearing NAB2^G221W^ mutations were outcompeted or acquired during final steps of progression such as a MAPK (MAP3K1^E224X^) mutation that potentially fostered the emergence of highly aggressive subclones. Presumably, the latter are capable of transitioning into the subarachnoid space and survive in the CSF. The comparative analysis of MBM and BMCs of patients 8, 27 and 35 validated a high genetic concordance (M1 vs BMC1-M1; 99.2%/ M4 vs BMC1-M4; 97.4%/ Pat 35 vs BMC2; 51.9%/ Pat 27 vs BMC3; 51.5%). Besides class I mutations, we discovered subclones carrying a class III BRAF mutation (BRAF^N581Y^) that were maintained in the tumor (Pat35) and *in vitro* (BMC2) next to NRAS^G12C^ mutant clones.

Moreover, we identified a minor subclone in Pat8/M4 carrying a probably damaging low- frequent NOTCH3^S1128P^ mutation (AF=0.03), (Supplementary Fig. 10b) that was maintained by *in vitro* cell culture conditions (BMC1-M4, AF=0.46; Supplementary Fig. 10c, Supplementary Table 6). Overall, these results suggest that the general composition of genetic subclones was not affected by the *in vivo*-to *in vitro* transition. Nonetheless, a minority of clones that was suppressed *in vivo* emerged *in vitro*.

### BMCs exhibited unique responses towards therapeutic drugs

The development of progressive intracranial disease is frequently observed in melanoma patients and the consequence of drug resistance. Next, we ascertained whether and to which extent the drug resistance conferring mechanisms of patient’s tumors were conserved in BMCs. Patient’s 8/M4 and 38 exhibited mutations in BRAF (BRAF^V600K^; AF=0.91) and RAC1 (RAC1^P29S^; AF=0.36) but lacked resistance-mediating secondary mutations in BRAF or acquired mutations in MEK1^98^ or NRAS^99^. We tested the sensitivity of BMCs towards low (1nM-10nM) and high doses (30nM -10 µM) of dabrafenib, one of the first-line modalities for patients with BRAF^V600E/K^ MBM. As expected, TargetSeq and pyrosequencing (Fig. 6b, Supplementary Fig. 10d) confirmed the persistence of BRAF^V600E/K^ mutated tumor cells *in vitro*. Remarkably, BMC1-M4 cells showed a significant response to low doses (3nM, 10nM) of dabrafenib (IC_50_=7.65 nM). In contrast, BMC1-M1 cells originating from a drug-naïve tumor were refractory to low doses (IC_50_=224.4 nM) of dabrafenib (Fig. 6c, first and second panel). This finding likely reflected the loss of resistance at least partly due to the switch from BRAFi to ICi in the therapeutic course of Pat8. BMC4 cells were refractory to low doses of dabrafenib (IC_50_=47.72nM) (Fig. 6c, third panel and Supplementary Fig. 11a). Congruently, BMC2 cells carrying a BRAF^N581Y^ mutation showed no response to dabrafenib (Supplementary Fig. 11b, left panel). Although previous studies suggested that the expression of NGFR/CD271 drives resistance to BRAFi^77,100^, we observed no correlation of CD271/NGFR levels with response to dabrafenib.

The evolution of BRAFi/MEKi therapy-resistant genetic subclones that hamper drug response and consequentially drive metastatic progression is frequently observed in melanoma patients. Although chemotherapeutics show limited efficacy in disseminated melanoma and are mostly applied for palliative care, they might be effective in BRAFi- or ICi- resistant tumors. We assessed the efficacy of paclitaxel, an actin filament interfering drug in BMCs that exhibited a low response towards dabrafenib. Paclitaxel was effective in intrinsic dabrafenib-resistant BMC4 cells (IC_50_=153.3 pM) or less-sensitive BMC1-M1 cells (IC_50_=114.5 pM, Fig. 6d, first and second panels, Supplementary Fig. 11b, right panel). In addition, the frequent expression of the MET receptor tyrosine kinase in BM of lung and breast cancer suggests a clinical relevance of small molecular inhibitors of MET for the treatment of BM^101, 102^. We discovered that the MET receptor was among the Ecad^high^ subset enriched genes (FClog2=2.1, p=3e-03) and significantly higher expressed in MBM than EM (p=2.7e-05; Supplementary Fig. 12a, left and center panels). Moreover, MET expression was significantly higher in the Ecad^high^ subset of MBM (n=43, p=1.4e-04, two-way anova) than EM (n=50, p=0.41; Supplementary Fig. 12a, right panel). We evaluated the expression and activation of MET in MBM and observed activated MET signaling (phosphorylation at Tyr1234/1235, p-MET^Tyr1234/1235^) in a subset of tumors (Supplementary Fig. 12b, upper panels). Fluorescence in-situ hybridization revealed that MET expression was not linked to gene amplification (Supplementary Fig. 12b, lower panel and 12c upper panel). In addition, TargetSeq confirmed absence of MET receptor mutations in our MBM cohort. MET receptor signaling is a well-known driver of cell migration and associated with activity of small Ras-related GTPases such as RAC1 which in turn trigger the formation of cortical actin-containing membrane ruffles and lamellopodia^103–105^. The expression of MET was maintained in BMCs and localized at membranes including ruffles (Supplementary Fig. 12c, lower panels). Therefore, we expected that highly migratory cells are sensitive to the non-ATP competitive MET-inhibitor tivantinib (ARQ197), which has been clinically approved for hepatocellular carcinoma (NCT01755767) and lung cancer (NCT01395758) among other non-melanoma related studies. We discovered that BMCs responded to ARQ197 (BMC1-M1: IC_50_= 351.8 nM; BMC1-M4: IC_50_=376.1 nM; BMC2: IC_50_= 596.3 nM; BMC4: IC_50_= 291.4 nM) irrespective of the degree of dabrafenib response. However, we observed a higher sensitivity in conventional melanoma cell lines (A375: IC_50_= 200.8 nM, A2058: IC_50_= 242.2 nM and MeWo: IC_50_= 106.1 nM; Fig. 6d-e, Supplementary Fig. 12d).

Taken together, the comparative analysis of longitudinal MBM and concordant cell lines provided insight into the persistence and stability of genetic subclones and suggests that low-passage BMCs reflect the genetic landscape of tumors and serve as *in vitro* model systems.

### The loss of Ecad expression recapitulates steps of *in vivo* progression

Supposedly, the first metastatic dissemination to local sites such as regional lymph nodes selects for a subset of cells establishing the metastatic cascade. A prerequisite for the latter process is the survival of circulating cells in the blood stream and lymphatic vessels and the capability of adaptation to microenvironmental cues, prevailing at distant organ sites^106^. Like extracellular niches, the intracerebral tumor cell niche is built up by stromal cells such as astrocytes, microglia and infiltrating immune cells (Fig. 7a, “*in vivo*”) that release growth factors and exosomes in response to their interaction with tumor cells. Although we observed an overall concordance of MBM and BMCs, the derivation of BMC1-M1 cells (Fig. 7a, “*in vitro*”), was accompanied by differential expression of 362 genes (p=5.3e-04), which was likely triggered by the *in vivo*-to-*in vitro* switch (Fig. 7b). Particularly, we found a loss of the Ecad^+^ state (Fig. 7c, left panel) of the initial tumor (M1; FClog2=-6.775, p=6.41E-05). Nevertheless, a minor Ecad^+^ subfraction was maintained *in vitro* (Fig. 7c, right panels), in line with previous findings suggesting that Ecad expression is unstable in cell lines^107^. Next, we sought whether the *in vitro* loss of Ecad^+^ cells reflected mechanisms of *in vivo*progression, hence the derivation of the therapy-resistant subclone M4 from M1. Comparative analysis of whole transcriptome data of Pat 8/M1 vs. M4 and their respective tumor-derived cell lines (BMC1-M1; BMC1-M4) revealed a clear proliferative-to-invasive switch of MBM cells but concordance of M4 and BMC1-M4 cells (Fig. 7d). Apart from NGFR, we focused on differentially regulated transcription factors in our survey and identified significantly decreased levels of STAT5A (FClog2=-2.468, p=3.88e-02) and MITF (FClog2=-2.840, p=2.16e-02) in early and late passage BMC1-M1 cells. Levels of FOSL1 (FClog2=8.145, p=1.39e-05), FOSL2 (FClog2=3.277, p=1.29e-02) and POU3F2 (FClog2=3.280, p=1.36e-02) expression were increased. Likely, the latter factors are responsible for the activation and maintenance of cell autonomous, hence niche-independent programs. We hypothesized that both the emergence of progressive subclones such as M4 and the *in vitro* transition of M1 cells were driven by a comparable set of molecular programs and likely triggered by changes of niche-provided factors. The comparative analysis of genes differentially regulated between M1 vs. BMC1-M1 and M1 vs. M4 revealed a common set of ∼100 (termed “progressive”) genes that promoted the conversion of Ecad^+^ into CD271^+^ MBM cells *in vivo* and *in vitro* (Fig. 7d, right panel) among them NGFR, FOSL1 and FOSL2 (Supplementary Table 7). As STAT5A has been associated with Ecad expression^108^, we surveyed the correlation of STAT5A and Ecad in MBM and extracranial metastases of melanoma (TCGA-MM). STAT5A but not STAT5B was correlated with Ecad expression in MBM (R=0.837; p=1.00e-04, Fig. 7e and Supplementary Fig. 13a, left panel) and among the upregulated genes (FClog2=1.581, p=1.04E-05) in the Ecad^high^ subset. In addition, STAT5A was expressed in a subset of Ecad^high^ extracranial metastases (n=315, EM; p=2e-04, Fig. 7e) but not in brain metastases (n=6, BM) or primary tumors (n=151, PT) of TCGA-MM. FOSL1 was correlated with the CD271 status of TCGA-MM (EM, PT; Supplementary Fig. 13a, right panel), however FOSL expression was not associated with CD271/NGFR expression in MBM (Supplementary Table 4). Next, we assessed the distribution and activation of STAT5A and expression levels of Ecad in CD271^high^ and low MBM.

**Fig. 7:**
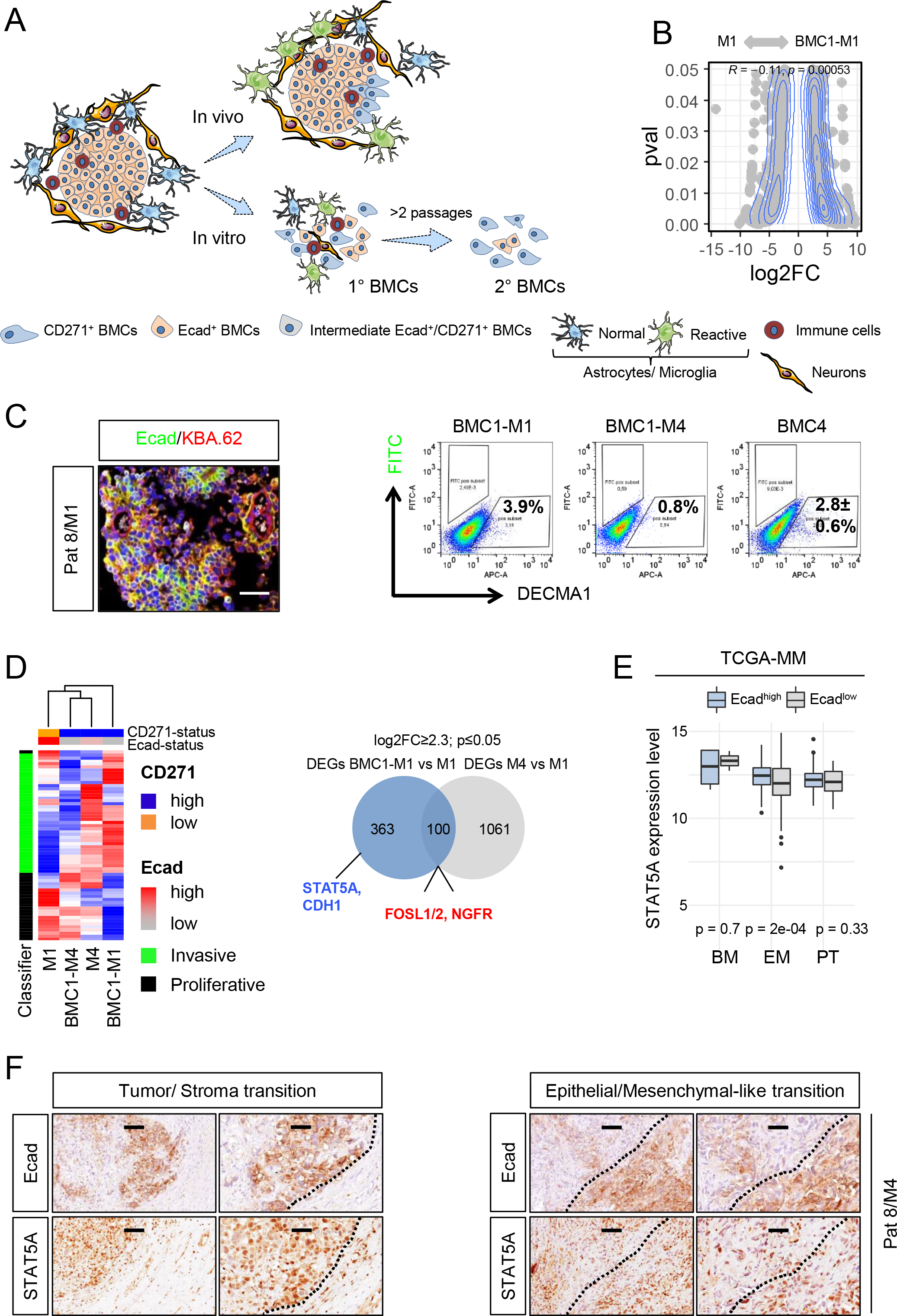
The brain microenvironment maintains an Ecad^+^ tumor cell phenotype. a.) Schematic representation of tumor progression *in vivo* and phenotype-switch *in vitro*. *In vivo*: MBM are surrounded by normal and reactive astrocytes/microglia, neurons and immune cells. The interaction of tumor cells with microenvironmental cells determines tumor progression stages. Likely, inflammatory processes foster MBM progression. *In vitro*: the derivation and maintenance of tumor cells was associated with a loss of microenvironmental cells and transition of Ecad^+^ into CD271^+^ cells via transient Ecad^+^/CD271^+^ cells. b.) Scatterplot representation of M1 and BMC1-M1 enriched genes. c.) IF of M1 for Ecad and KBA.62 (left panel) and analysis of cell surface levels of Ecad by flow cytometry and DECMA1 antibody; 50,000 cells were recorded (right panels). d.) Supervised heat map depicting levels of invasive and proliferative classifier genes in M1, M4 and associated BMCs. Ecad and CD271 status is color-coded (left panel). Venn-diagram displays a comparison of differentially regulated genes (DEGs) between M1 vs. BMC1-M1 and M4 vs. M1, 100 genes common to both groups were identified. NGFR, FOSL1 and FOSL2 were among the significantly, most up-regulated genes. STAT5A and CDH1 were expressed in M1 but downregulated in BMC1-M1. e.) Box plots represent a significant association of levels of STAT5A and expression of Ecad in extracranial metastases (EM, p=2e-04) but not BM or primary tumors (PT, p=0.028) as determined by two-way anova using TCGA-SKCM (melanoma, MM) data. f.) IHC of M4 shows overlapping expression of Ecad and STAT5A and low STAT5A levels in brain parenchymal cells (left panels) and reduced levels of STAT5A at Ecad^high^-to-Ecad^low^ transition sites. Bars indicate 50 µm.

Immunohistochemistry analysis for Ecad and total STAT5A revealed that even CD271^high^ tumors such as M4 comprised Ecad^+^ cells that were likely maintained in progressive tumors and not converted into CD271^+^ cells reflecting at least two different co-existing cellular stages (Supplementary Fig. 13b, upper scheme). Ecad^+^ expressing cells exhibited activated STAT5A signaling, hence nuclear STAT5A and a reduced infiltration into adjacent tissue (Fig. 7f, left panels). Moreover, we identified a decreased nuclear localization or expression of STAT5A at epithelial-like into mesenchymal-like transition sites (Fig. 7f, right panels). However, intersecting areas of Ecad^neg^/STAT5A^+^ and Ecad^+^/STAT5A^neg^ tumor cells suggested that STAT5A activation was required but probably not sufficient for driving Ecad expression. Considering the high plasticity of melanoma cells, Ecad and CD271 are likely interconnected. To address whether Ecad^+^ evolved from CD271^+^ cells or vice versa (Supplementary Fig. 13b, upper scheme), we established a transcriptional dual-reporter system facilitating the tracing of the Ecad^+^ subset via Ecad promoter-controlled expression of RFP. We monitored the NGFR/CD271 subset via a 3’-UTR-GFP reporter that enabled the detection of NGFR-mRNA stability (Supplementary Fig. 13b, lower scheme)^34, 109, 110^. The additional constitutive expression of iRFP enabled the general labeling of reporter cells independent from phenotype switching processes (Supplementary Fig. 13c, upper panels). As CD271/NGFR-GFP^+^ cells are generally sustained *in vitro*^109^, we traced FACS-enriched RFP/iRFP cells (Supplementary Fig. 13c, lower panels). The initial (100%) Ecad^+^ fraction was decreased by 38.9±13.6% (p ≤ 0.01) 2 days after the FACS-based isolation (Supplementary Fig. 13d), suggesting that Ecad^+^ subsets are unstable and not sustained by standard 2D *in vitro* conditions. However, live cell-imaging revealed rare (<0.1%) derivation of Ecad^RFP+^ cells from NGFR^GFP+^ (Supplementary Fig. 13e and movies 1 and 2), or double negative cells.

Next, we investigated whether forced STAT5A expression was sufficient to restore Ecad expression and increased the Ecad^+^ subset in BMCs. Therefore, BMC4 cells were transduced with wildtype (STAT5A^wt-GFP^), mutant (STAT5A^S710F-GFP^) STAT5A or empty control plasmids (Supplementary Fig. 14a, left panels). STAT5A^S710F-GFP^ represents a constitutive active form with enhanced tetramer forming capacity^111^. We observed a marginal but significantly increased number of Ecad/DECMA1^+^ cells in STAT5A^S710F-GFP^ (3.6±0.8%, p=0.053) but not in STAT5A^wt-GFP^ (1.4±0.3%) or control cells (1.6±0.6%; Supplementary Fig. 14a, right panel), suggesting that STAT5A activation might be required but was not sufficient to generate or maintain an Ecad^+^ cell phenotype.

### CD271 cooperates with a network of progressive-genes to mediate cell migration and invasion

Considering that CD271 controls cell migration and MBM progression, we performed whole transcriptome profiling of BMC1-M1 with a highly efficient inducible knockdown of NGFR. We tested the functional reliability of shRNA in conventional (A375, WM35) and MBM-derived cell lines ∼7-14 days after doxycycline (DOX)-treatment (Supplementary Fig. 15a). We -1.13; p ≤ 0.05) down-regulated genes in BMC1-M1. Moreover, we identified 125 CD271-dependent genes that were commonly downregulated in BMC1-M1 cells and in a lymph node metastasis-derived cell line (T2002, GSE52456, Supplementary Fig. 15b). Among them were SOX4, a master-regulator of EMT^112, 113^ and PTPRZ1 a mediator of stemness in glioblastoma^114^. As both cell lines were unrelated and comprised different sets of mutations (T2002, BRAF/NRAS^wt^; BMC1-M1, BRAF^V600E^), the CD271-associated regulation of genes was probably context independent. The EMT-promoting transcription factor SOX4 (FClog2=-2.9; p=1.1E-17, Supplementary Table 8) was among the most significantly downregulated genes in BMC1-M1 and T2002 cells.

The expression of CD271 was correlated with a suspension phenotype of melanoma cells^84^ that was frequently observed in BMCs. Suspension cells (SCs) were viable and their serial reseeding established adherently growing CD271^+^ cells (Fig. 8a, upper panel) which in turn subsequently shed suspension cells. The phenotype was more evident in BMC1-M4 (10.3±0.7%) than in BMC1-M1 (3.6±2.2%) cells (Fig. 8a, lower panel) and was significantly reduced (3.3±1.8%, p=0.013) by the knockdown of CD271 (Fig. 8a, lower panel).

**Fig. 8:**
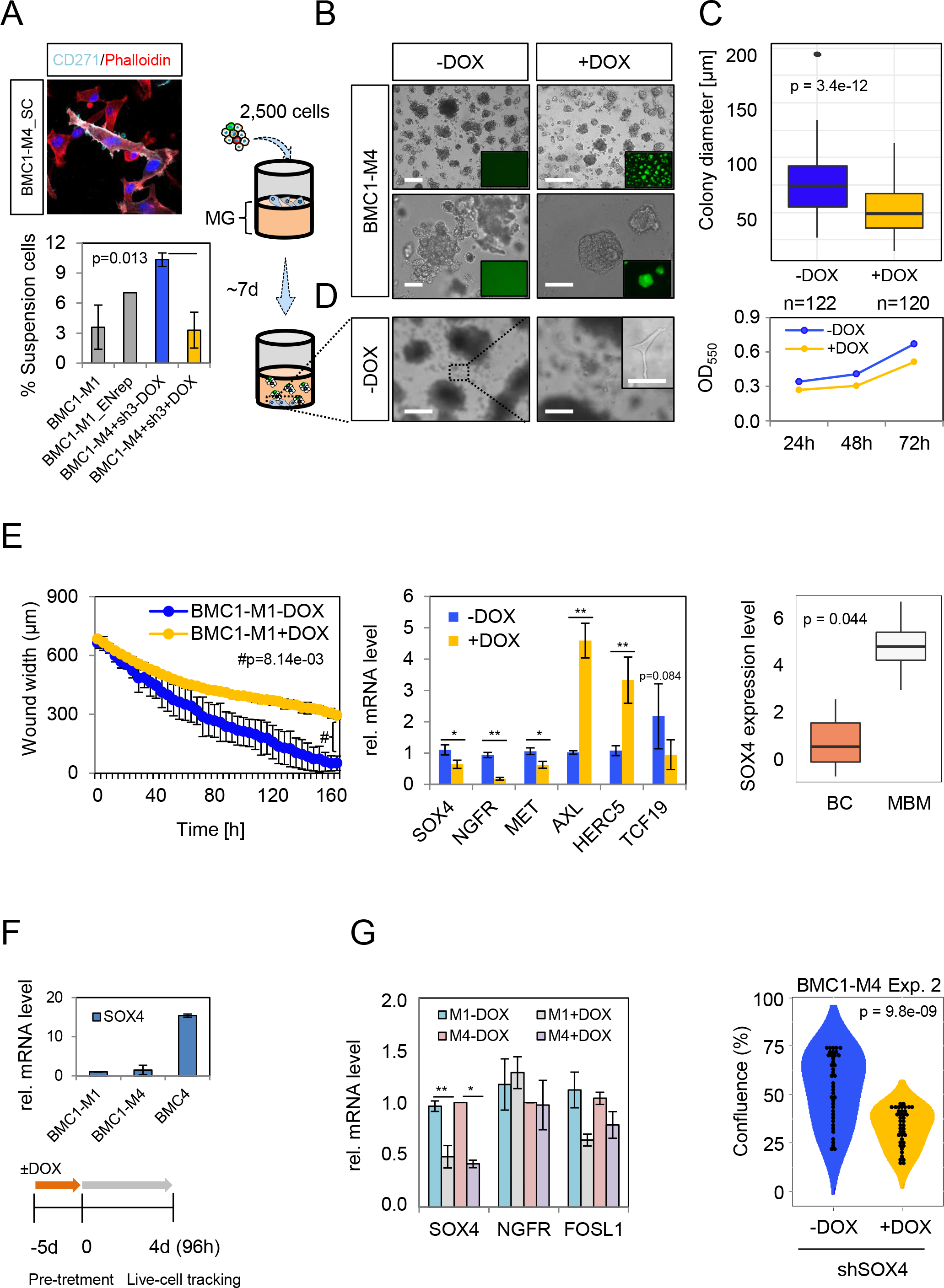
CD271 and SOX4 control migratory and proliferative properties of BMCs. a.) IF of re-seeded suspension cells (SC) of BMC1-M4 cells for CD271 and phalloidin shows the maintenance of CD271 expression of SCs after re-attachment (upper panel). Bar diagrams indicates the percentage of suspension cells relative to adherent cells of BMC1-M1 and BMC1-M1 reporter cells (Ecad/NGFRrep) and BMC1-M4 in absence (-DOX) and presence (+DOX). The latter significantly decreased the number of SCs (p=0.013). The seeding of 2,500 CD271 knockdown and control BMC1-M4 cells on a solidified layer of matrigel enabled the investigation of cell invasion (right scheme). b.) The knockdown of CD271 significantly reduced spheroid formation (p=3.4e-12) and extracellular matrix invasion c.) Quantification of changes in spheroid growth as determined by colony diameters (µm), n=122 (-DOX) or n=120 (+DOX) spheroids were analyzed (upper panel). Two-dimensional proliferation was not significantly affected (lower panel). d.) Invasive cells that crossed the matrigel layer and attached to the bottom of wells were only observed in absence of DOX. Insets show GFP expression upon DOX treatment, bars indicate 50 µm. e.) Live cell-imaging based scratch migration assay of BMC1-M1 cells revealed a significant decrease (p=8.14e-03) in the migratory capacity and a delayed wound closure (left panel). qPCR for potential drivers of migration in BMC1-M4 cells. The knockdown of CD271 was accompanied by reduced levels of SOX4, and MET but increased expression of AXL and HERC5 (center panel). Box plot shows a significant higher expression of SOX4 in MBM (n=16) than brain controls (BC, n=3), p=0.044 (right panel). f.) qPCR illustrates levels of SOX4 in indicated BMCs (upper panel), experimental scheme of DOX treatment of BMC1-M4 and BMC4 cells prior to live cell- imaging based assessment of proliferation for 4d. g.) the efficient knockdown of SOX4 was validated by qPCR (left panel) and led to a strong significant (p=9.8e-09) decrease in proliferation (right panel). Significance was determined by a two-tailed paired t-test. Values depict mean±sdv, *p < 0.05, **p < 0.001.

Following, we assessed whether the modulation of CD271 levels affected invasion and 3D- spheroid formation. We then seeded 2.5x10^3^ cells of either control cells (-DOX) or cells with a validated knockdown of CD271 (+DOX) on a matrigel layer (Fig. 8b, left scheme). Control cells invaded the matrigel layer and formed spheroids after ∼7 days that in turn shed cells into the matrigel (Fig. 8b, left panels). The concomitant expression of GFP (Fig. 8b, right panels; inlaid) enabled the tracing of cells upon DOX induction (Fig. 8b, right panels).

Downregulation of NGFR expression significantly (p=3.4e-12) reduced the diameter of spheroids (Fig. 8c, upper panel, Supplementary Fig. 15c) and affected the two-dimensional growth (Fig. 8c, lower panel). A minority of cells that likely featured a highly invasive phenotype completely crossed the matrigel layer and attached to the vesseĺs bottom (Fig. 8d). This process was not observed upon NGFR knockdown. Finally, we assessed whether decreased expression of CD271 was sufficient for hampering migratory processes of BMCs. The migratory phenotype of BMC1-M1 and BMC2 cells, both featuring different types of BRAF mutations was significantly reduced upon NGFR downregulation with no obvious difference in the extent of inhibition (Fig. 8e, left panel, Supplementary Fig. 15d). In line, we observed reduced levels of drivers of EMT/ cell migration such as SOX4^115^, MET receptor (MET)^116^, and TCF19^117^ and increased expression of AXL (Fig. 8e, center panel) in CD271 knockdown cells. SOX4 likely acts as a master regulator of EMT^118^ and was expressed in all MBM investigated (FClog2=4.44±0.97; range: 2.6-6.3) but expressed at lower levels in BC (FClog2=0.45±1.64; range: -1.04-2.21; p=0.044) (Fig. 8e, right panel). Moreover, SOX4 expression levels were comparable in primary and metastatic melanoma (TCGA set; data not shown) and sustained in BMCs (Fig. 8f, upper panel) suggesting a general role of SOX4 in the maintenance of melanoma cells *in vivo* and *in vitro*. To dissect the role of SOX4, we performed a SOX4 knockdown in BMC1-M4 and BMC4 cells and tracked changes in proliferation of knockdown (+DOX) and control cells (-DOX) via live-cell imaging for 5 days (Fig. 8f, scheme). We observed sustained NGFR and FOSL1 expression after 10d of DOX treatment in cells with a significant knockdown of SOX4 (Fig. 8g, left panel) as well as a significant decrease in proliferation of BMC1-M4 (p=9.8e-09; p=5.7e-03) and BMC4 (p=1.3e- 05; p=1.1e-04) (Fig. 8g, right panel; Supplementary Fig. 15e). Therefore, we suggest that SOX4 acts downstream of CD271 to control properties of MBM-derived cells and likely maintains the proliferative and migratory state of MBM.

## Discussion

The emergence of brain metastases is frequently observed in patients with melanoma, lung and breast cancer^119^ presenting a devastating step in the course of disease. Despite ongoing progress, the long-lasting therapeutic control of BM remains challenging and the majority of patients exhibit a poor prognosis due to progressive and multiple BM^16^. However, the emergence of multiple BM is poorly understood and might involve (i) the establishment of a founder clone that gives rise to multiple subclones or (ii) the transition of dormant micrometastases into actively proliferating macrometastases triggered by microenvironmental cues and/or therapeutic interventions^3, 119, 120^. Whether primary tumors can directly metastasize to the brain or tumor cells need a “priming” step while acquiring the capability of crossing the BBB during the sequential process of metastasis, remains elusive.

Here, we performed a molecular, genetic and epigenetic profiling of BM of stage IV melanoma (MBM) patients and uncovered that the expression of the epithelial marker Ecad and NGFR subdivided MBM into drug-naïve, potentially responsive and drug-resistant, progressive tumors. Moreover, we established and characterized a panel of stable MBM-derived cell lines that were molecularly distinct from conventional melanoma cell lines and enabled the analysis of programs establishing cellular dependencies. In addition, we provided evidence that therapeutic interventions inhibiting the acquired constitutive activation of BRAF likely foster the phenotype switch from Ecad^+^ into NGFR/CD271^+^ cells. Considering that the expression of NGFR in melanoma cells accompanies a network of associated genes such as mediators of a NCSC-phenotype^34, 59, 121^ - which was recently identified as a major driver of MRD^35^ - we suggest that the upregulation of NGFR and associated factors such as FOSL1 hallmarks the fate of relapse in the brain and intracranial progression. Although brain metastasis *per se* is associated with poor prognosis of melanoma patients, high levels of NGFR likely accelerate progression potentially via secreted extracellular vesicles^37^. Indeed, CD271^high^ MBM exhibited an invasive and stem-like phenotype predominantly observed in extensively treated patientś tumors. Therefore, CD271/NGFR^+^ microenvironmental cells potentially facilitate tumor cell invasion and migration or aid in maintaining micrometastasis.

Although the molecular programs that control subclonal evolution and development of multiple brain metastases remain elusive, the analysis of longitudinal metastases of one of our patients (Pat 8) provided insights into the genetic landscapes. We observed that mutations in BRAF or NRAS, RAC1, MYC, and CARD11 defined the genetic landscapes of tumors. However, TargetSeq revealed that every tumor carried private mutations that were mostly maintained in MBM-derived cell lines. The successful establishment of stable, expandable tumor-derived cell lines served as an important tool for the investigation of mechanisms of drug-response and provided insights into cellular dependencies and the persistence of known driver mutations such as of BRAF, NRAS, RAC1 and genetic subclones. Though, the cultivation of tumor-derived cell lines and subsequent passaging is associated with global changes in methylome profiles^122^ and likely establishes cellular dependencies that are distinct from *in vivo* processes. We observed that BMCs differentially responded to dabrafenib, irrespective of presence of constitutive active RAC1 (RAC1^P29S^). BMC2, carrying a BRAF^N581Y^ mutation showed no response, as expected. BMC1-M1, BMC1- M4 and BMC4 carried a comparable ground state of BRAF- and RAC1- but not MEK1- or NRAS-mutations and responded differentially to dabrafenib. Interestingly, BMC1-M4 exhibited a higher sensitivity to dabrafenib than the correlated BMC1-M1 cells, which likely reflected the therapeutic course. Whereas Pat8/M1 received radiation therapy and was surgically removed prior to systemic dabrafenib/trametinib (BRAFi/MEKi) therapy, BMC1-M4 emerged under BRAFi/MEKi therapy. The excision of M4 tumor occurred after completion of BRAFi/MEKi and subsequent ICi (nivolumab/ipilimumab) therapy. This therapeutic switch was potentially causative for adoption of a dabrafenib sensitive phenotype of M4 that was sustained *in vitro*. However, we cannot exclude that both tumors M1 and M4 independently emerged from pre-existing dormant micrometastases that stem from a common extracranial tumor and therefore carried a common set of ground-state mutations. However, both tumors might have acquired additional mutations that have not been picked up by TargetSeq due to limitations of the amplicon panel. As resistance to BRAFi is frequently observed in melanoma patients^123, 124^, we sought for additional potential therapeutics. We discovered that MET receptor signaling was activated in MBM. Consistently, all BMCs responded towards the clinically approved non-ATP competitive inhibitor ARQ197 (tivantinib)^125^. Moreover, the actin filament interfering drug paclitaxel was effective in BMCs, suggesting that both therapeutics are potentially effective in BRAFi resistant MBM. Cell lines such as BMC1-M1 and BMC1-M4 reflected the phenotypes of the initial tumors and maintained the patterns of genetic clones that were found in initial tumors. Moreover, we observed rather an enrichment than a depletion of genetic subclones.

However, the Ecad^+^ phenotype of MBM (M1)-derived cells was not maintained *in vitro* and during progression *in vivo* and was likely promoted by serum supplied TGFβ or inflammatory processes^126–128^. Hence, specialized serum-free, chemically defined media may support the maintenance of Ecad^+^ states *in vitro*. In addition, TGFβ signaling inhibits the activation of STAT5 signaling and antagonizes STAT5-mediated gene expression^129, 130^ and might thus have reduced the effect of STAT5A expression and restoration of an Ecad^+^ phenotype in BMCs. The loss of STAT5A signaling therefore might act in concert with increased TGFβ signaling to support the progression of MBM *in vivo* and phenotype switching *in vitro*. The latter possibly involves FOSL1 and STAT5A expression controlling the transition of Ecad^high^/CD271^low^ into Ecad^low^/CD271^high^ MBM, hence defining cellular subsets of MBM. Additional factors such as SOX4 control melanoma cell proliferation. Recently, Ecad was identified as a survival factor in invasive ductal carcinomas during different steps of metastasis^131^ and might therefore control similar processes during extracranial and intracranial metastasis of melanoma.

Molecular programs that drive the emergence and progression of solitary and multiple brain metastases are poorly understood. Unraveling the mechanisms that control the migratory and invasive properties of tumor cells and drive intracranial spreading might leverage the development of new therapeutic strategies preventing intracranial progression. Here, we identified Ecad and NGFR/CD271 associated networks that potentially determine progression stages and drug response of MBM.

## Data Availability

All data produced in the present study are available upon reasonable request to the authors under accession numbers EGAS00001005976 and EGAS00001005975

https://ega-archive.org/

## Supplementary figure legends

**Supplementary figure 1**: **Molecular subsets of post- and pre-MAPKi treatment melanoma.** a.) Whole tumor scans of extensively treated (coded) MBM (Pat 15, 17) indicating high levels of CD271 expression; scale bars indicate 5 mm or 50 µm. b.) Dot-plot representation indicates significant difference in levels of E-cadherin (CDH1) but not melanocyte-specific markers such as dopachrome-tautomerase (DCT) and MLANA (melanA; melanoma antigen recognized by T cells 1) in pre- versus pos-treatment melanoma (study GSE77940). c.) GSEA of pre- versus post-treatment melanoma for representation of gene signatures specifying CD271-associated genes (CD271-core UP) or MAPKi-induced EMT

**Supplementary figure 2**: **Genetic characteristics of MBM.** a.) Supervised clustering of MB and brain controls (Cortex, Pons, Cerebellum; Cereb.) demonstrating a clear difference and the presence of molecular signatures (Classifier) specifying the presence of exosomes, melanosomes, focal adhesion or extracellular matrix (ECM). b.) TargetSeq of MBM and brain controls (K1, K2) using a 50 gene panel (CHP2v) revealed mutations (single nucleotide variants, SNV) in 11 genes and provided insight into copy number changes (CNV gain, loss), insertion and deletions (INDEL) and nonsense mutations (STOP-gain). c.) Types of BRAF and NRAS mutations identified in (a), the mutation status was not determined (ND) in 10 MBM (21%).

**Supplementary figure 3**: **Methylation of the ITGB7 promoter is associated with improved survival.** a.) Determination of tumor cell content via expression of PRAME (%, related to BMC1-M1 cells, left panel). Scatter plot indicating the similarity of BRAF^mut^ and BRAF^wt^ MBM regarding the degree of methylation of ∼850k CpG islands in promoter regions (right panel). Red dots indicate the most significant differentially methylated CpGs (FDR-adjusted p-value<0.05). b.) Presentation of 46 differentially methylated CpG islands within promoters of 14 genes identified by global methylation analysis of BRAF^mut^ and BRAF^wt^ -values (mean, β≤0.5, unmethylated; β≥0.5, methylated CpGs.) c.) Survival analysis of melanoma patients based on levels of expression of ITGB7 in brain metastases (n=80, EGAS00001003672) showing that high ITGB7 levels are associated with favorable outcome.

**Supplementary figure 4**: **The expression of NGFR discriminates M1 and M4.** a.) MRI, H&E and CD271 staining of M2, M3 reflecting differences in location and phenotypes of subclones and stages of progression. b.) Scatter plot of ∼13,000 genes of M1 and M4 indicated the difference of both tumors (R=0.85, p<2.2e-16, two-tailed paired t-test). c.) IF of a lymph node metastasis (LN-MET, M1 concordant) for single staining of Ecad (extracellular domain) and CD271/p75^NTR^. d-e.) IF for CD271, KBA.62 and Ecad/DECMA1 (N-terminal domain) of M1 indicates the rarity of CD271^+^ cells. In c-e, DAPI served as nuclear stain, confocal imaging was performed. Scale bars indicate 50 µm.

**Supplementary figure 5**: **E-cadherin was expressed among different melanoma progression stages.** a.) Box plot representation of levels of Ecad/CDH1 in brain metastases (BM, n=79) and non-concordant extracranial metastases (EM, n=59) of study EGAS00001003672. Statistical testing (*two-tailed paired t-test) revealed no significant difference (p=0.61) of Ecad levels (left panel). Box plot representation of levels of Ecad/CDH1 in primary and metastatic melanoma of the SKCM (n=472, TCGA) set including rare (n=6) BM. A one-way anova (^) indicated the significant difference (p=0.0013) of Ecad levels among tumors (right panel). b.) Representation of levels of Ecad expression (log2 FPKM) in MBM samples (blue bars) and brain controls (orange) and MBM-derived cell lines (BMC1-M1 and BMC1-M4). c.) Log2FPKM levels of NGFR expression in melanoma samples. d.) Box plots depict that levels of Ecad or NGFR significantly subdivide MBM into subgroups, (Ecad^high^ vs. low, p=3.5e-07; CD271/NGFR^high^ vs. low, p=1.1.e-06), right panels. e.) Enrichment of MITF-target signature genes in Ecad^high^ MBM of study EGAS00001003672 (n=80, left panel) and MBM of this study (n=16, center panel). No enrichment of MITF-target genes was observed in the CD271/NGFR^high^ subgroup of MBM (rigt panel). The analysis was performed with 10,000 permutations.

**Supplementary figure 6**: **Characterization of CD271^high^ and TIL^high^ subsets of MBM.** a.) Box plots depict a significant association of expression levels of NGFR with BRAF^wt^ (p=0.014) and expression of CDH1 with BRAF^mut^ (p=0.013) MBM. Single-sample GSEA (ssGSEA) depicts that nine signatures among them signatures associated with p75^NTR^/NGFR signaling and brain metastasis in breast cancer significantly (p<0.05, adjusted by Benjamini-Hochberg) subdivides MBM into CD271/NGFR^high^ and low subsets (color coded). c.) GSEA of CD271^high^ vs. low MBM shows a significant enrichment of signature genes that are predictive for breast cancer brain metastasis (NES=1.727, FDR-adjusted p-value = 0.003, left panel). IHC for CD3, a pan-marker of T lymphocytes subdivides MBM into TIL^high^ and low (center panel) and shows immune cell infiltration in M4 (right panel). Bars indicate 50 µm. d.) Box plots indicate that expressions levels (log2 FPKM) of CD3D (p=2.8e-07) and CD8A (p=2.6e-04) significantly subdivided MBM into TIL^high^ and low subsets (left panels) and are of prognostic relevance as depicted by Kaplan-Meier survival curves. Analyses were performed with expression and survival data of study EGAS00001003672, higher levels of CD3G (p=0.06) and CD8A (p=0.0029) were associated with increased survival (right panels). e.) GSEA of TIL^high^ and low subsets revealed increased inflammatory response (NES=2.449, FDR-adjusted pval<0.001) and astrocyte activation (astrogliosis, NES=1.842, FDR-adjusted pval<0.001) in TIL^high^ tumors. GSEA was performed with 10,000 permutations. In (a) and (d) a two-tailed paired t-test was performed.

**Supplementary figure 7**. **AXL and CD271 expression in BMCs.** a-b.) Flow cytometry of adherently grown BMC2, BMC3 and BMC53 for levels of CD271 or suspension cells (BMC2). c.) Box plots indicate that levels of AXL expression (log2FPKM) between BC and MBM are not significantly different (p=0.061, two-tailed paired t-test). d.) IF and confocal imaging depicted intracellular and membrane localization of AXL in BMC1-M1 and BMC1-M4 cells (left and center panels) and revealed AXL expression of in a MBM (Pat 4), right panel. e.) Co-expression and localization of CD271 (green) and p-AXL (red) in Pat 15. AXL phoshorylation was rarely observed in MBM (Pat 15, right panels). Bars indicate 50 µm.

**Supplementary figure 8**: **MBM-derived cell lines (BMCs) show distinct migratory capacities and transcriptomic profiles.** a.) Content of proliferative, Ki67^+^ cells in a drug-naïve (M1) and therapy-resistant (M4) MBM and associated *in vitro* cell culture models (left panels) and capacity of BrdU incorporation (right panel) indicate the maintenance of a proliferative phenotype of M4-derived (BMC1-M4) cells *in vitro*. Ki67^+^ and BrdU^+^ cells (% mean±sdv) were determined by counting of stained cells of three independent experiments. b.) Live-cell imaging based scratch-wound assay depicting an increased migratory phenotype of BMC2, BMC1-M4 and BMC4 but not BMC1-M1 and BMC3 cells. Values depict changes in wound width (µm mean±sdv) of three independent experiments. c.) Bar diagram displays values of wound width as determined by live cell-imaging at 0, 24, 48 and 72h after wounding. The migratory capacity of BMCs led to a significant decrease in wound widths after 24h (left panel). Values depict mean±sdv of three independent experiments. Significance was determined by a two-tailed paired t-test, values depict mean±sdv, **p < 0.001; ***p < 0.0001 d.) Experimental set-up and timeline of MRI imaging of BMC1-M1 transplanted CD-1 nude mice (n=3). MRI images (T1) of contrasted animals were taken after every week for a total of 49days (D). Tumor growth was detectable after 35days (yellow, dotted line). e.) Tumor growth over time as depicted by tumor volume (mm^3^) of BMC1-M1 inoculated animals.

**Supplementary figure 9**: **BMCs and conventional melanoma cell lines feature common and distinct properties.** a.) Western blot of whole protein lysates of BMCs for levels of CD271/p75^NTR^, quantification was related to GAPDH. b.) IF for CD271, GFAP (left panels) or CD271 and KBA.62 (right panel) and confocal imaging of brain tumors established by intracranial injection of 1x10^3^ A375 cells ∼25 days after transplantation. Three-dimensional representation was performed from z-stacks using the Arivis tool. c.) Correlation heat map of MBM, BCs, BMCs, a patient-derived cell line establish from a lymph node metastasis (T2002) and conventional cell lines MeWo, A375 and human melanocytes depicts a discrete clustering of conventional cell lines and BMCs. d.) The RAC1^P29S^ mutation forces formation of membrane ruffles in BMC1-M1 cells^132^ and showed co-enrichment of actin filaments and CD271 as depicted by z-stacking based three-dimensional modeling of BMC1-M1 cells stained for CD271 and phalloidin (left panels). The formation of membrane ruffles was established in WM35 and A375 cells following lentiviral infection with GFP or plasmids expressing constitutive active RAC1^P29S^ (right panels).

**Supplementary figure 10**: **TargetSeq provided insight into subclonal heterogeneity.** a.) Representation of allele frequencies (color-coded) of mutations identified by TargetSeq of MBM and BMCs. b.) PolyPhen2 prediction of a NOTCH3^S1128P^ mutation that raised in BMC1- M4 cells. c.) Changes of allele frequencies of indicated mutations among longitudinal metastases, CSF and cell lines of Pat8 demonstrating the permissive character of BRAF, RAC1 and MYC *in vivo* and of CARD11, NOTCH3 and MAG *in vitro*. d.) Chromatogram of BMC4 cells depicting the detection of a BRAF^V600K^ mutation by pyrosequencing.

**Supplementary figure 11**: **BMCs feature different types of BRAF mutations that determine the response to dabrafenib. a**.) Representation of confluence values relative to untreated controls and sensitivity of BMCs (BMC1-M1, BMC1-M4, BMC4) carrying druggable (BRAF^V600E/K^) and b.) non-druggable (BRAF^N581Y^) mutations (BMC2) towards dabrafenib, first panel. Error bars are not shown in this representation. Dabrafenib concentration is indicated, DMSO served as treatment control. Second panel: Fit-curve based computation of IC50 values, comparison of dabrafenib sensitive BMC1-M4 and less sensitive BMCs.

**Supplementary figure 12**: **The c-MET tyrosine kinase receptor control cell survival in MBM.** a.) Volcano plot depicting genes that are expressed in Ecad^low^ or Ecad^high^ MBM, c- MET (MET) was identified as Ecad-associated gene. Only significantly regulated genes (p<0.05) are shown (left panel). Box plots indicate expression levels of MET in non-classified BM and EM as well as in Ecad^high^ and Ecad^low^ tumors. MET was significantly higher expressed in BM (p=2.7e-05, center panel) and associated with Ecad^high^ tumors (p=1.4e-04, two-way anova), right panel. b-c.) IHC of a representative set of MBM for total (MET) and activated/ phosphorylated (Tyr1234/1235, p-MET) levels of MET showing concordance in three out of four MBM (upper panels). Fluorescence-in situ hybridization (FISH) using a MET-specific and centromere-specific (CEP7) probe (lower panels). Quantification of MET copies revealed no amplification when MET copies were related to CEP7 (c.), upper panel). IF of BMC1-M1 cells revealed expression and a membrane ruffle-associated enrichment of MET receptor (lower panels). d.) Dose-response of indicated BMCs towards tivantinib (ARQ197) a non-ATP competitive MET inhibitor was effective in dabrafenib-resistant BMC2 cells. Drug-response is depicted as decrease in confluence over time, tivantinib was titrated, (1nM-10 µM). Error bars indicate mean±sdv, eight technical replicates were analyzed, one out of three independent experiments is shown.

**Supplementary figure 13**. **A transcription reporter live-cell imaging approach identified Ecad-to-NGFR transition states.** a.) Box plots represent a significant association of expression of CD271/NGFR and levels of FOSL1 in EM (p=8.2e-07) and PT (p=0.028) but not BM (left panel). Box plot representation of STAT5B expression in BM, EM and PT of TCGA-SKCM. Two-way anova (^) revealed no significant association of STAT5B levels and expression of Ecad. b.) Scheme depicting a proposed interconnected relationship of Ecad^+^, CD271^+^ and intermediate state BMCs (upper part). Linear plasmid maps of reporters enabling the indirect tracking of NGFR expression via a NGFR-specific 3’-UTR-sequence based regulation of GFP mRNA stability and expression or tracking of Ecad expression via Ecad-promoter controlled expression of RFP or general tracking of cells via constitutively expressed iRFP (lower schemes). c.) IF of reporter cells prior to sorting, depicting unique and co-expression of reporters (upper panels). Bars indicate 50 µm. Lower panel: Fluorescence-activated cell sorting (FACS) based isolation of iRFP^+^/RFP^+^ BMC1-M1 cells. d.) Quantification of Ecad^+^ cells 3d post FACS revealed a significant (two-tailed paired t-test) decreased level of Ecad^+^ cells values depict mean±sdv, **p < 0.001. e.) Snapshots of NGFR^+^ into Ecad^+^ transitioning BMC1-M1 reporter cells (Movie 1) at days 0, 4. In (c) and (e) bars indicate 50 µm and 200 µm.

**Supplementary figure 14**: **STAT5A expression and activation is not sufficient for restoration of Ecad^+^ phenotypes of BMCs.** a.) IF of BMC4 cells retrovirally infected with an empty-control plasmid or plasmids expressing wildtype (STAT5A^wt^) or constitutive active (STAT5A^S710F^) for levels and localization of STAT5A. Transgene expression was monitored via IRES-GFP. Bars indicate 50 µm (left panels). Flow cytometry for DECMA1^+^ cells revealed a weak increase of Ecad/DECMA1^+^ cells in GFP^+^ (exhibited transgene expression) and GFP^-^ (did not exhibit transgene expression). Values depict mean±sdv of two biological replicates.

**Supplementary figure 15**: **SOX4 controls survival-mediating programs downstream of CD271 in BMCs.** a.) Western blot shows the efficient knockdown of CD271 in conventional (A375, WM35) and MBM-derived cell lines in presence of DOX (4 µg/ml) administered for 7- 14d. Tubulin served as loading control (left panel). Volcano plot shows significantly DEGs (FDR-adjusted p<0.05) in BMC1-M1 cells with a validated knockdown of CD271. SOX4 (padj=1.61e-13), SNAI2 (padj=1.36e-21) and HERC5 (padj=3.33E-69) were among the most significantly regulated genes, besides NGFR (padj=3.36e-09) (right panel). b.) Comparative analysis of candidate mediators of survival in T2002 and BMC1-M1 cells. Most commonly downregulated genes are shown, values depict log2 fold change (FC) related to -DOX cells (left panel). c.) Box plots depict a significant reduction (p<2.2e-16) in colony diameters of BMC1-M4 spheroids following knockdown of NGFR in a second independent experiment. d.) The knockdown of CD271 decreased the migratory capacity of BMC2 cells as indicated by changes in wound width (µm), left panel and relative (Rel.) wound density (%), right panel. e.) knockdown of SOX4 in BMC4 cells significantly reduced the proliferative capacity of cells, two independent experiments of eight technical replicates are shown (left and center panels). The knockdown of SOX4 was independently validated in BMC1-M4 cells (right panel). Significance was determined by a two-tailed paired t-test. Values depict mean±sdv, *p < 0.05, **p < 0.001.

## Acknowledgments

This research was supported using resources of the VetCore Facility (VetImaging | VetBioBank) of the University of Veterinary Medicine Vienna, Austria. We gratefully thank Mascha Osang for the careful proofreading of the manuscript. We are indebted to Petra Matylewski for excellent technical assistance.

## Author contributions

JR, ES, RK, AV, TR performed experiments and wrote the manuscript; JO performed craniotomy and provided MBM; GA, BB, CS performed intracranial injections and MRT; ST performed methylation profiling; MM performed DNA sequencing; SH, PK performed immunohistological expertise and performed immunophenotyping; DW, FW provided BMC53 cells, sections and expertise; FH, SKR, FG provided infrastructure; KJ, TR performed data analysis. All authors have read and agreed to the published version of the manuscript.

