## Supplementary figures and images for "Decoding molecular programs in melanoma brain metastases"

### Supplementary Figure 1

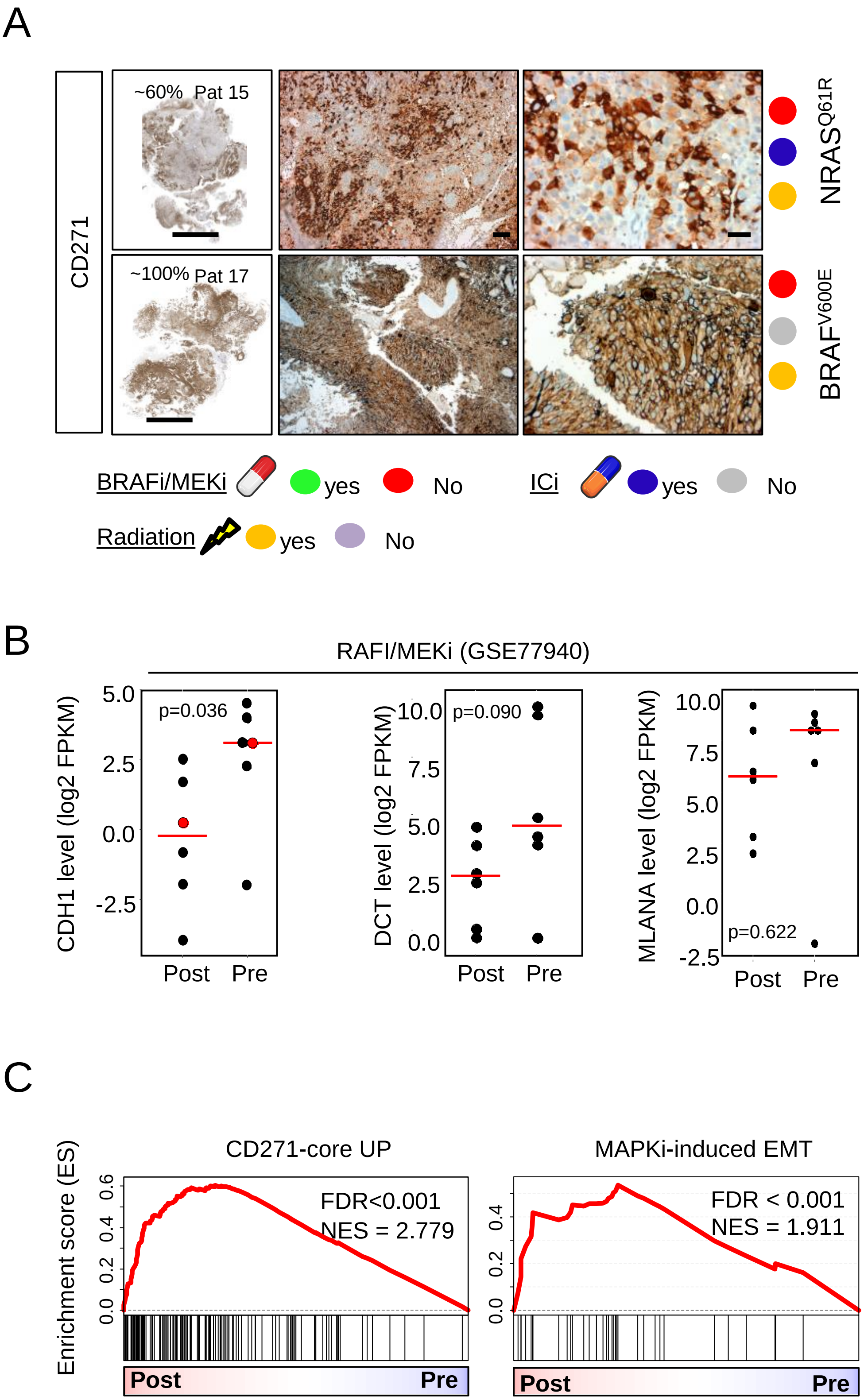

### Supplementary Figure 2

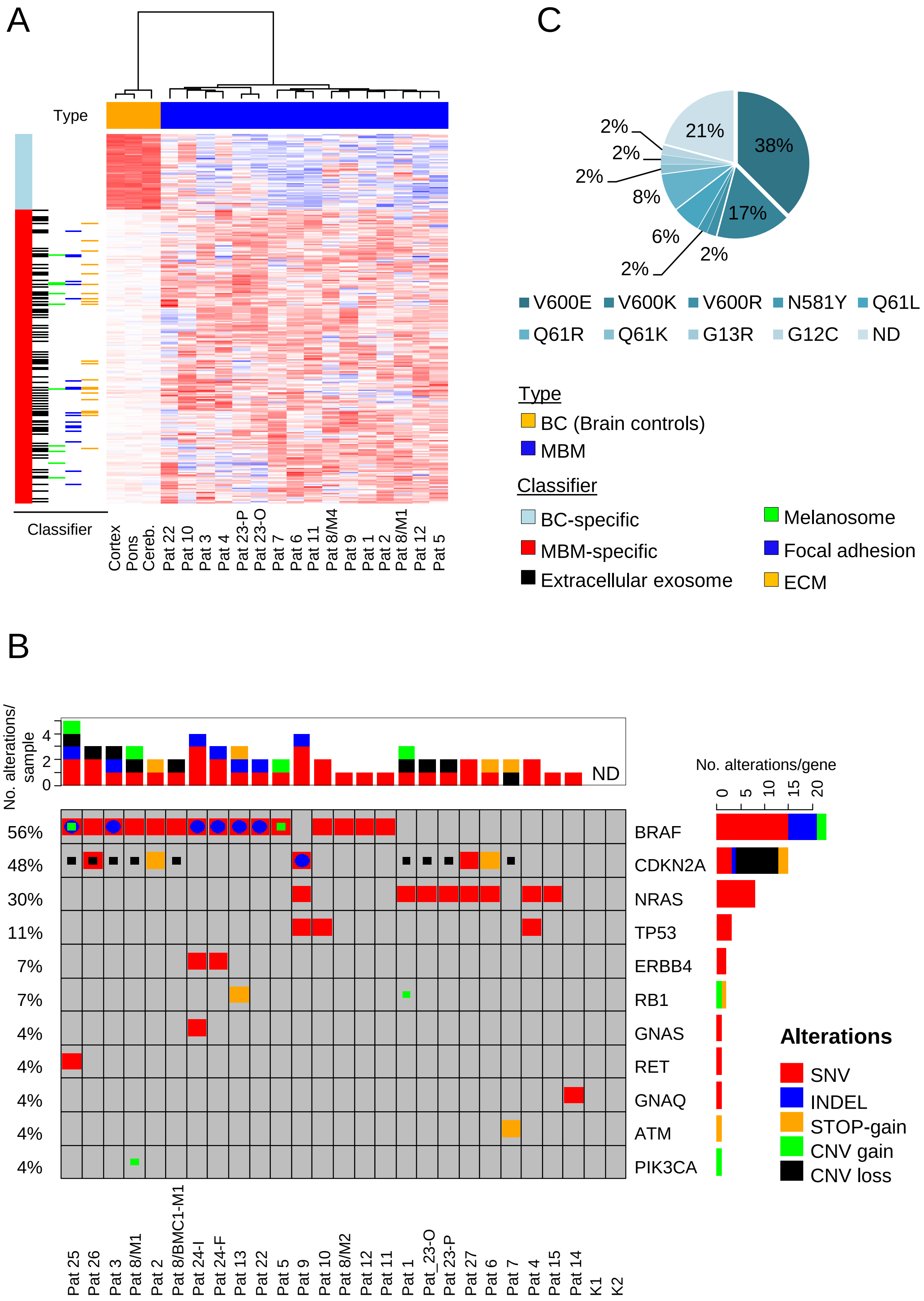

### Supplementary Figure 3

A

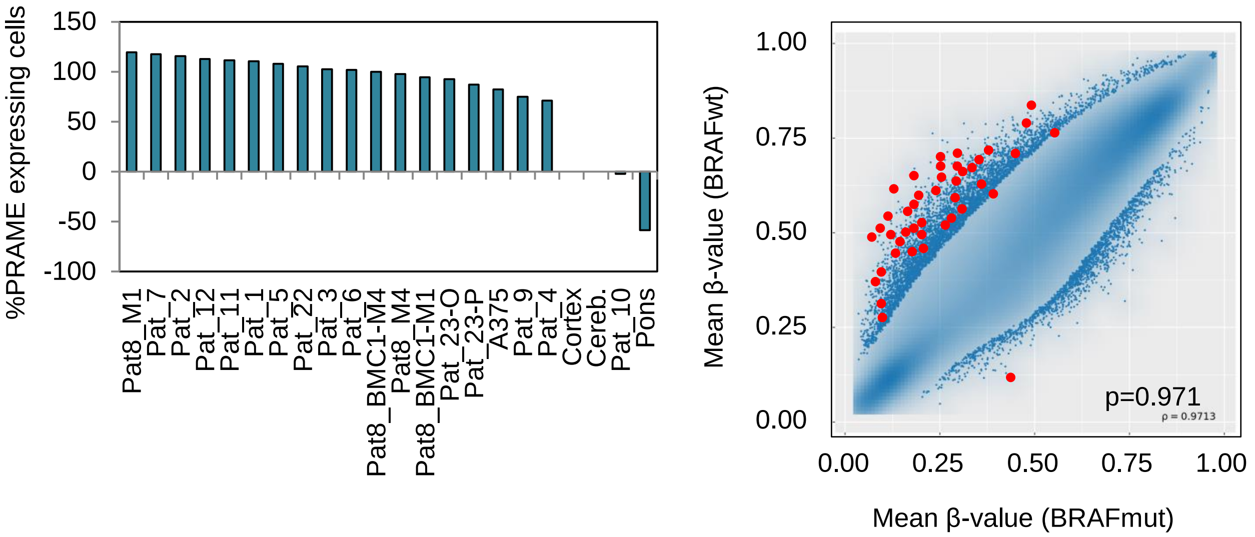


B

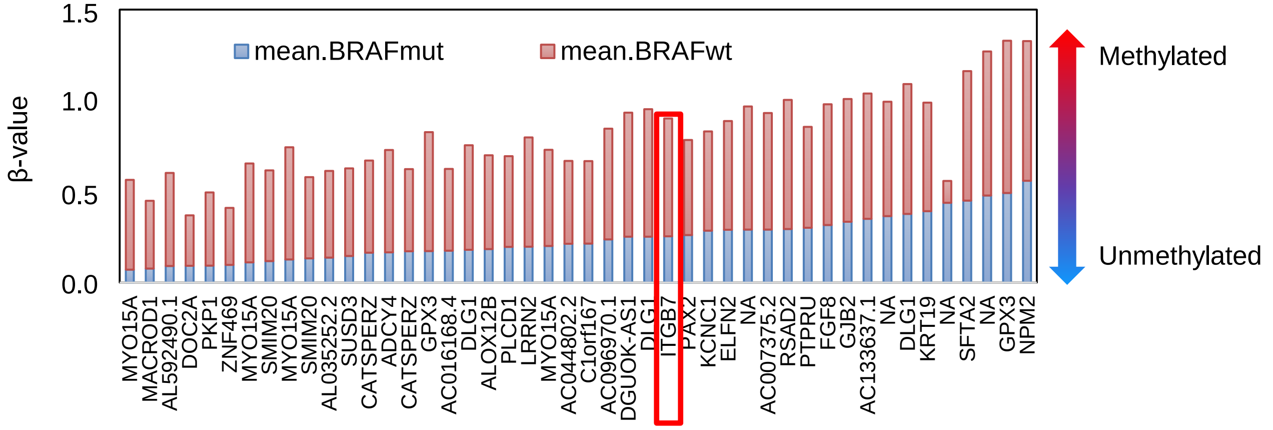


C

Survival analysis: brain metastases (n=80, EGAS00001003672)
featuring high or low mRNA levels of ITGB7

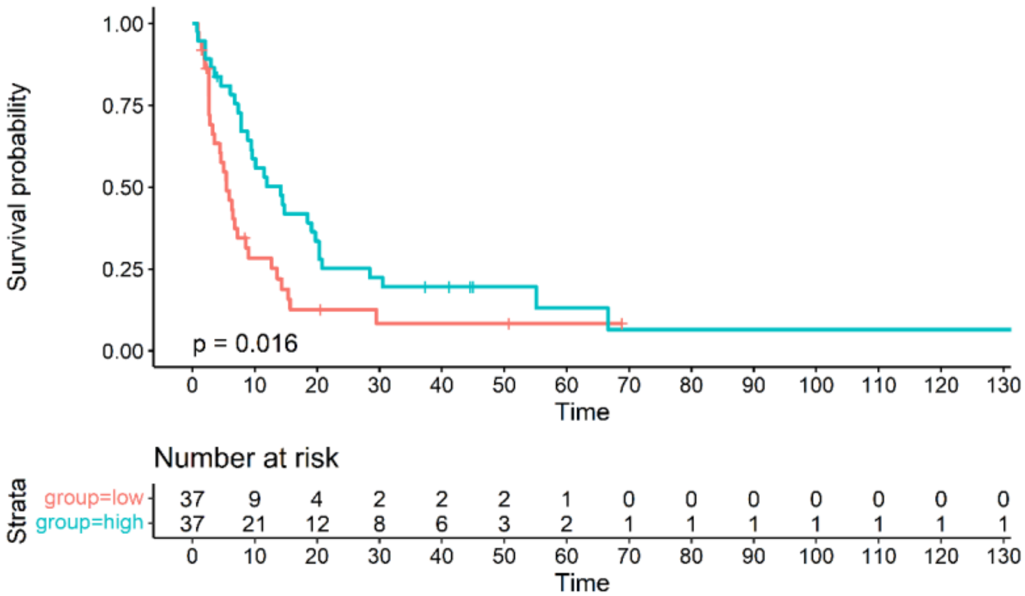

### Supplementary Figure 4

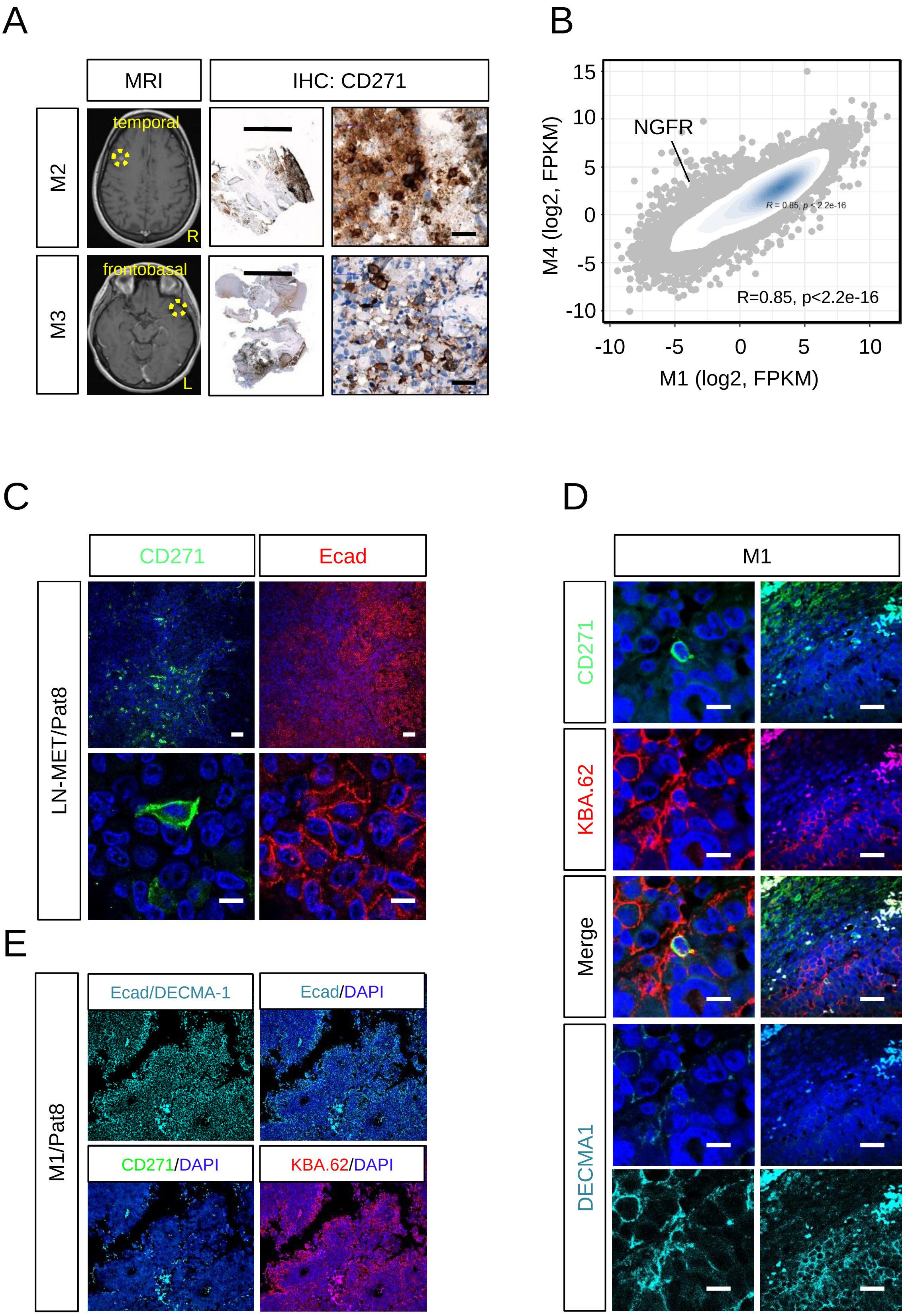

### Supplementary Figure 5

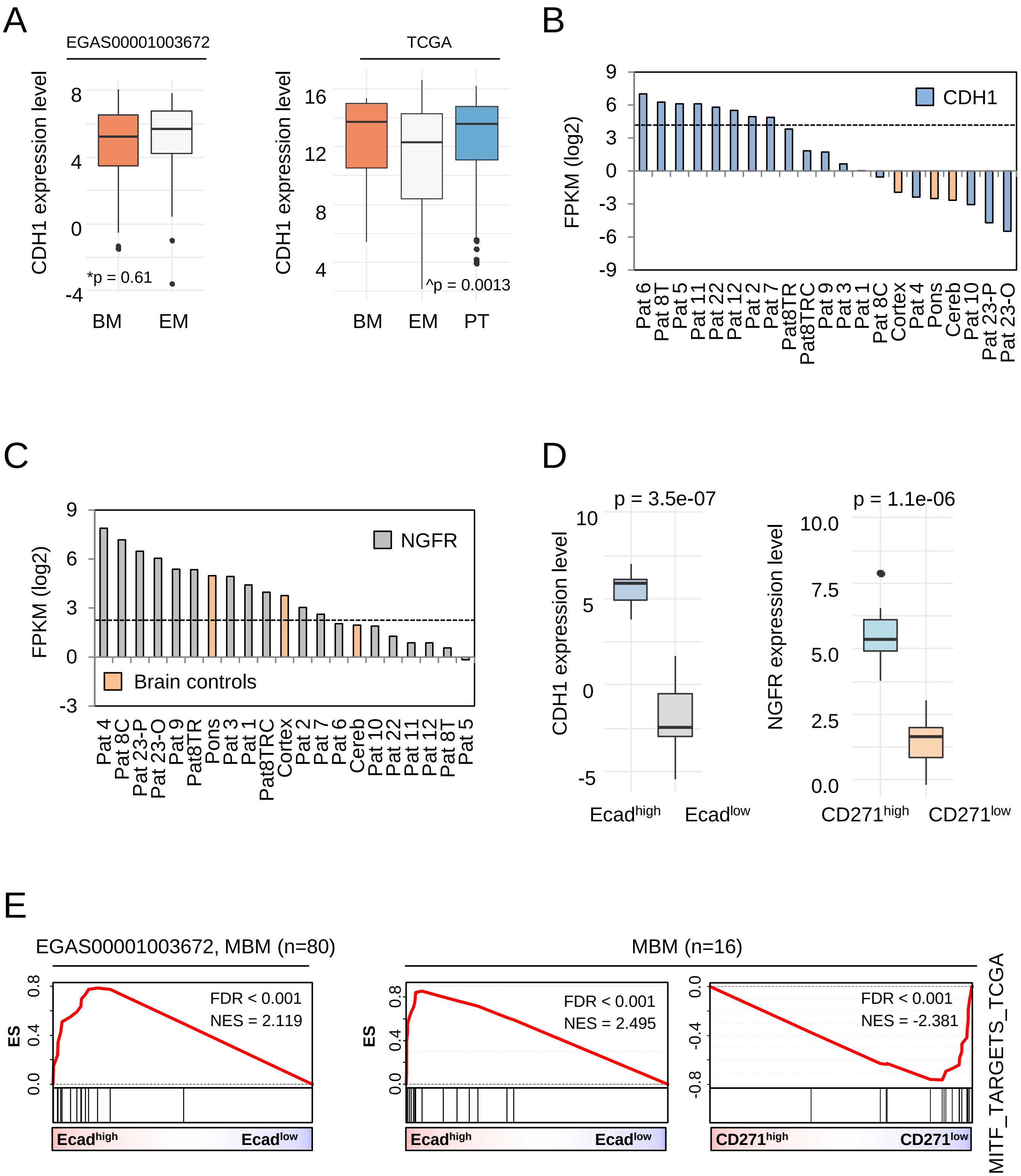

### Supplementary Figure 7

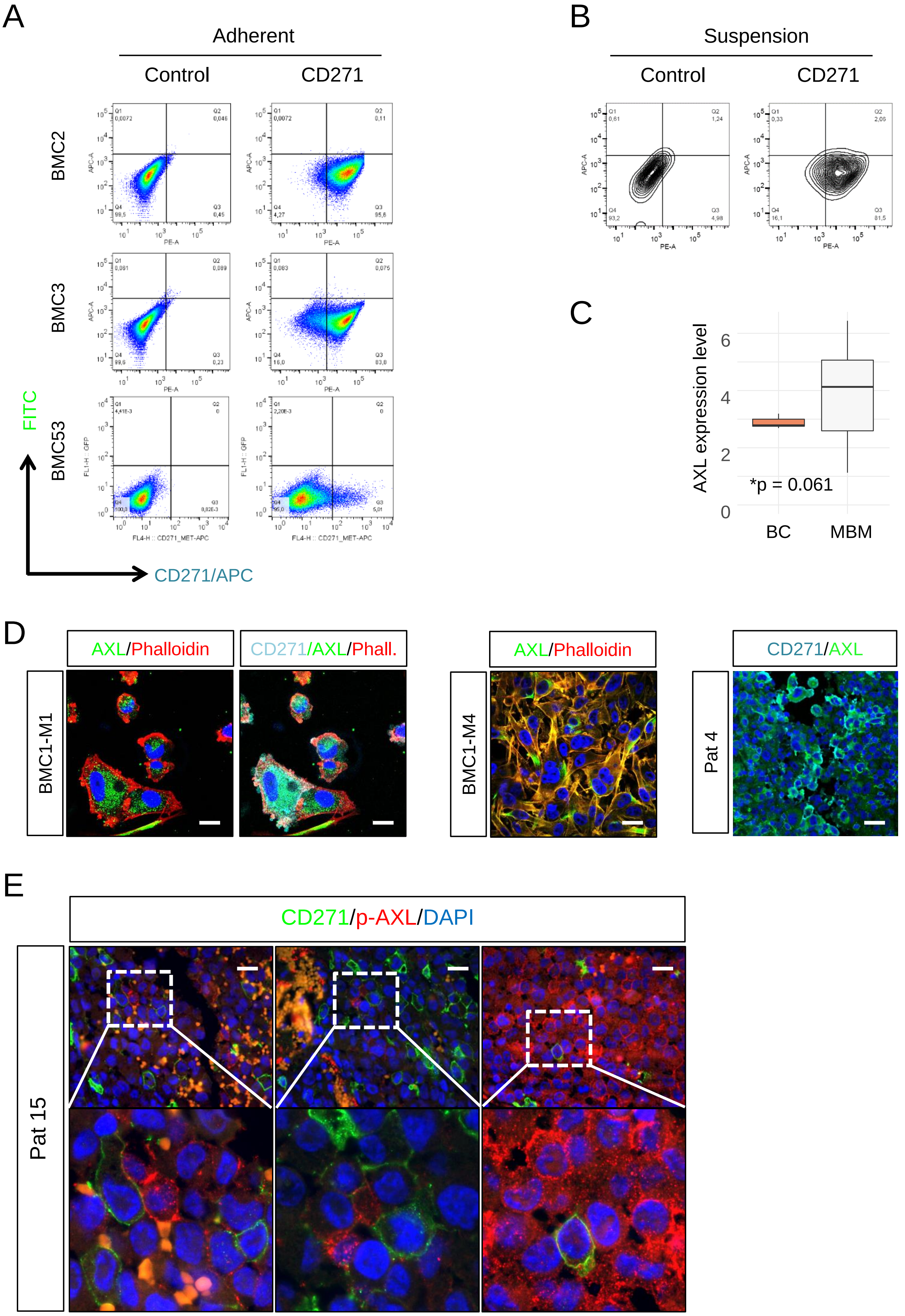

### Supplementary Figure 8

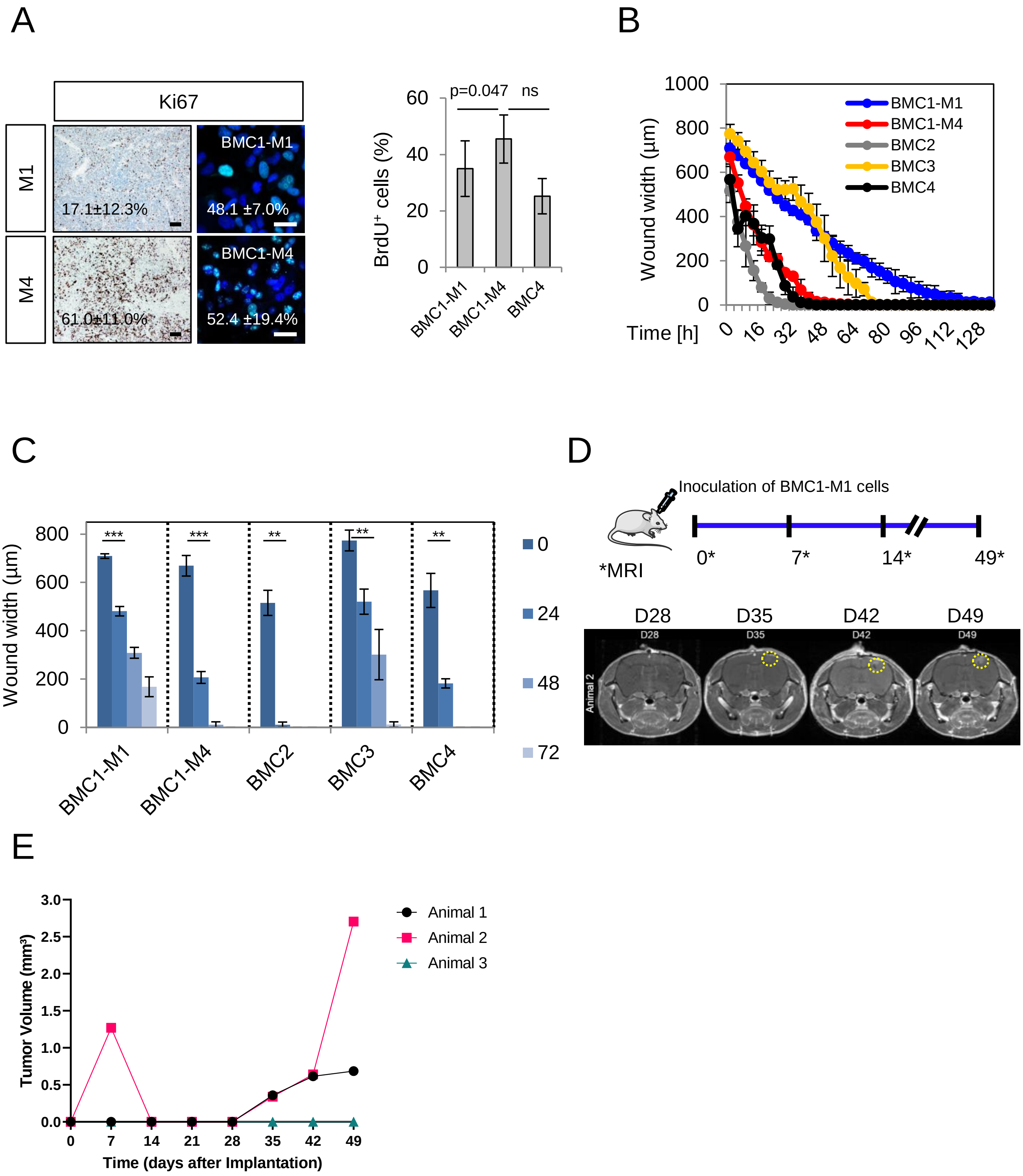

### Supplementary Figure 9

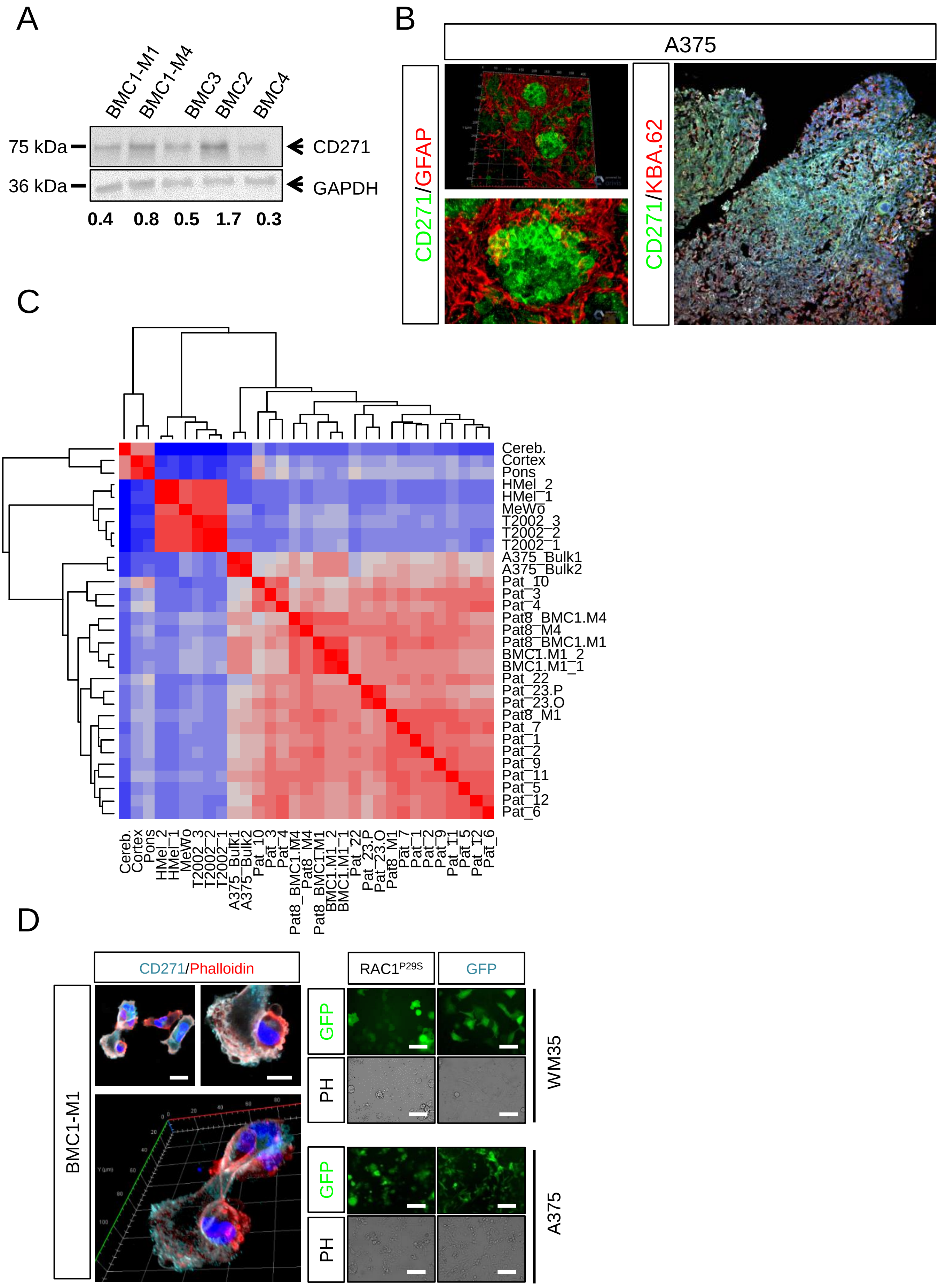

### Supplementary Figure 10

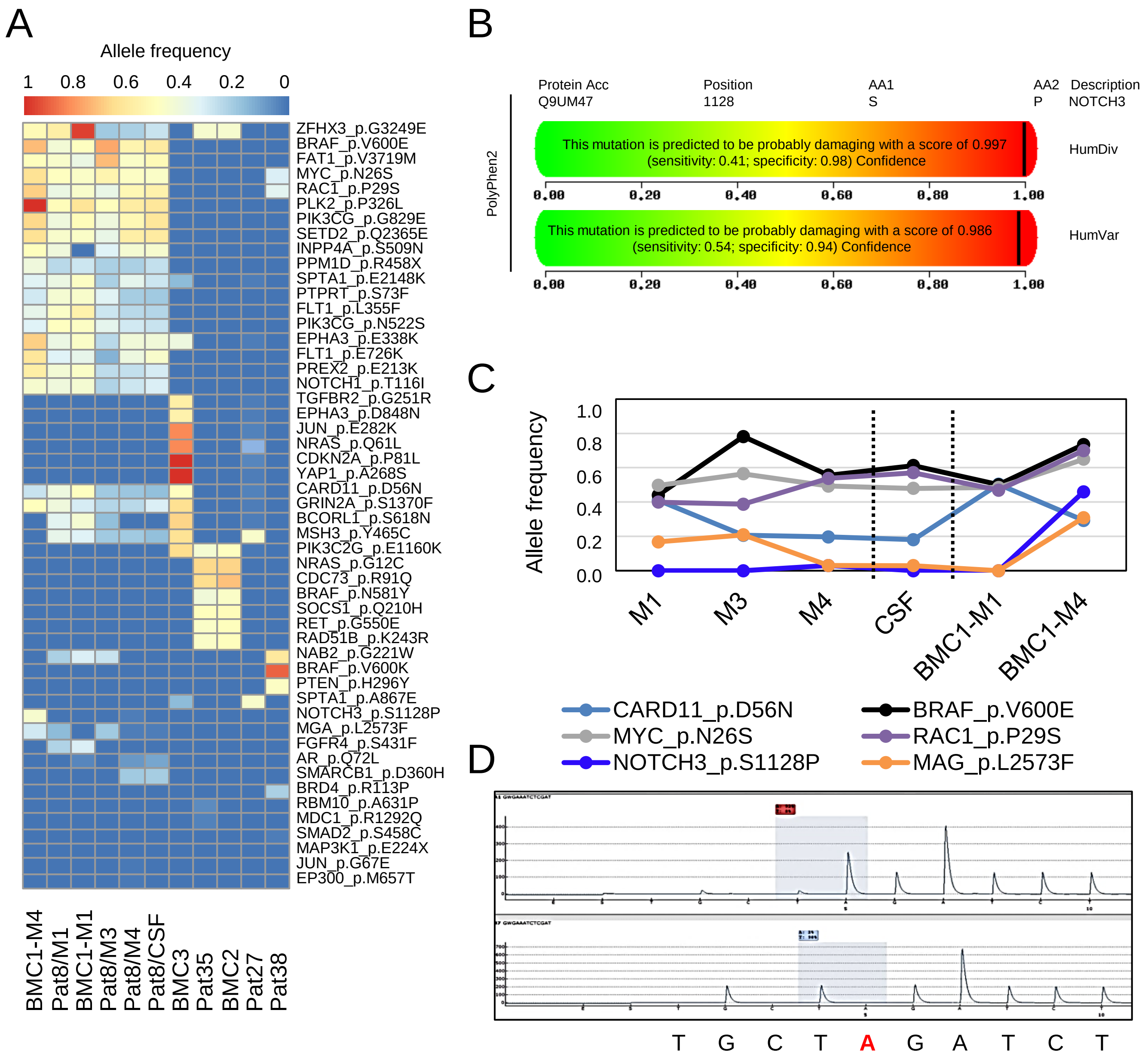

### Supplementary Figure 11

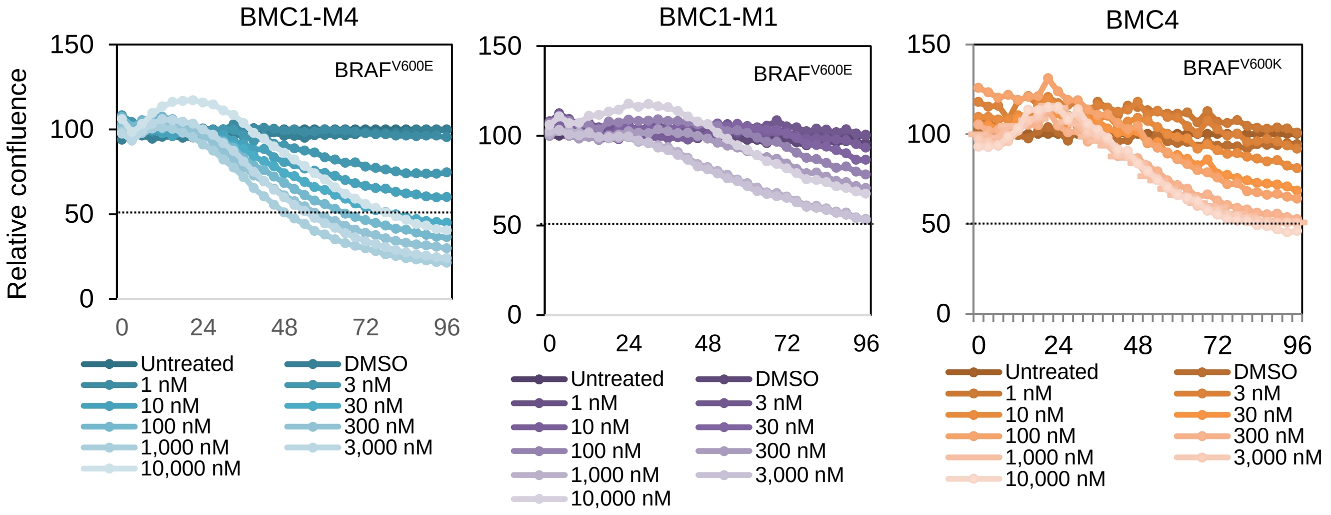

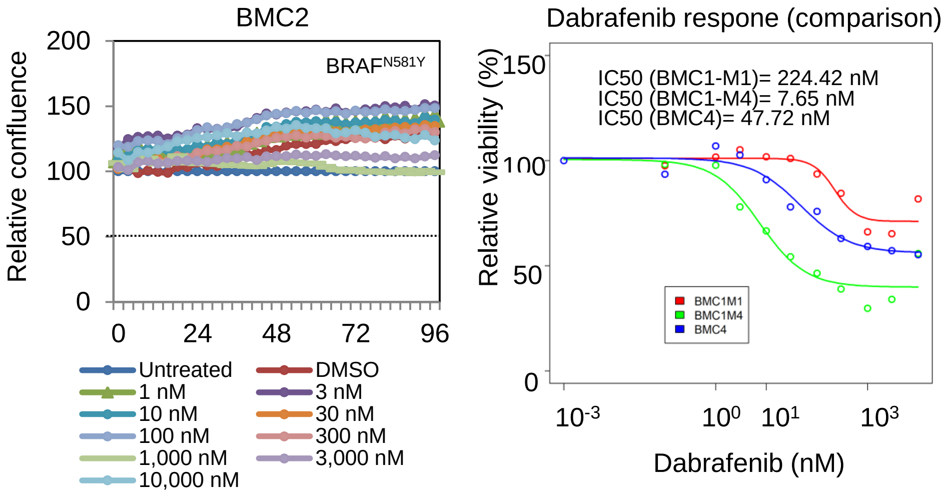

### Supplementary Figure 12

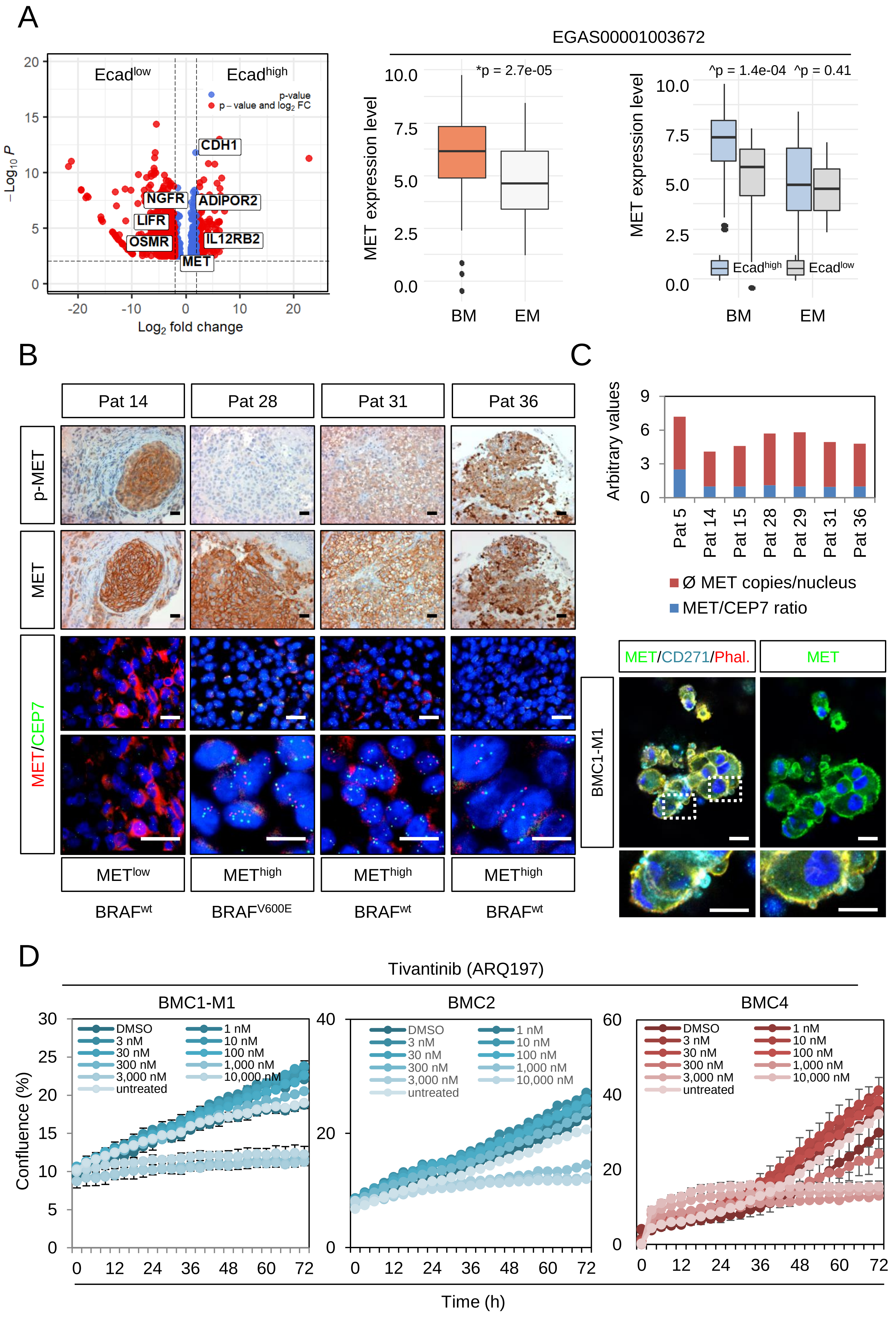

### Supplementary Figure 13

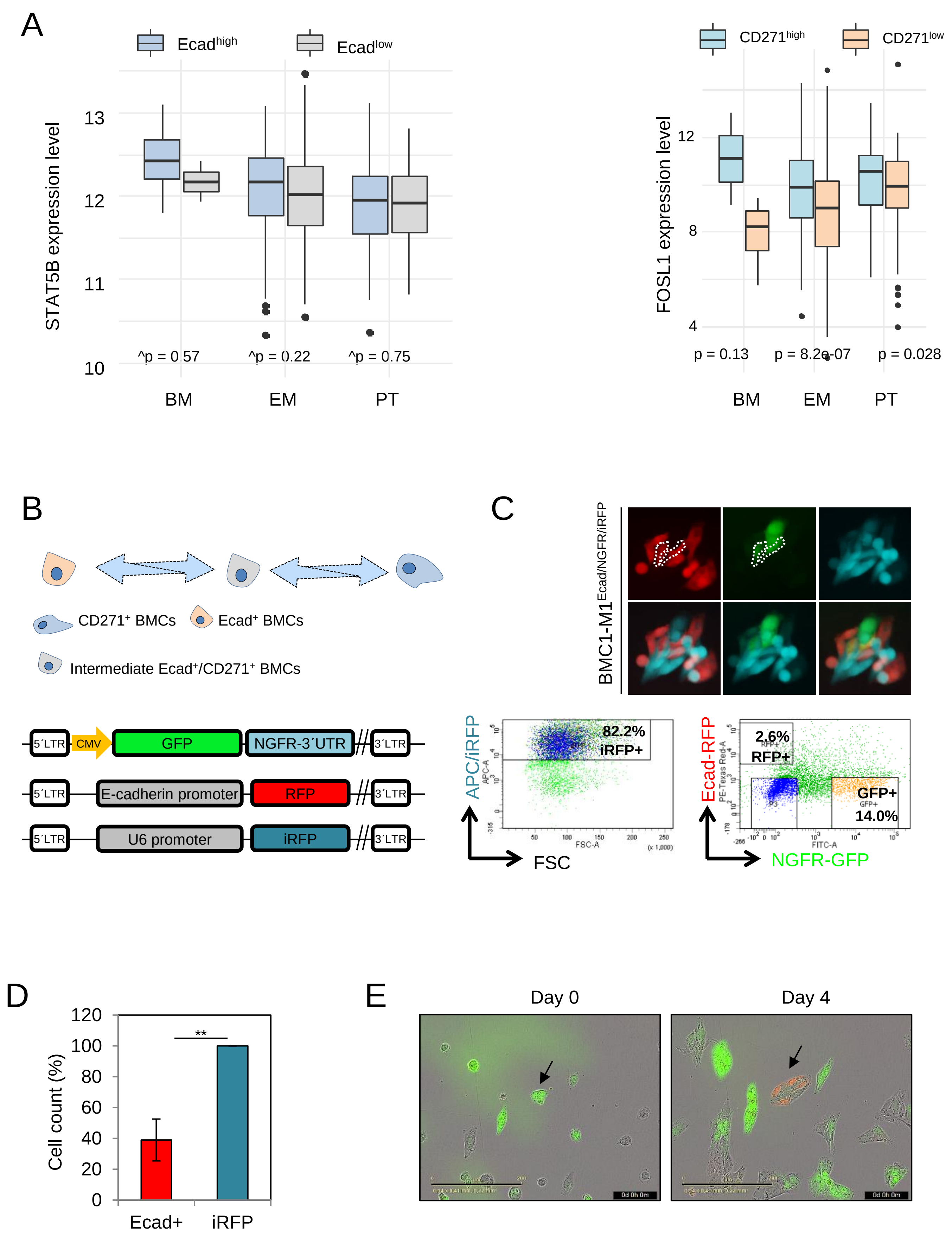

### Supplementary Figure 14

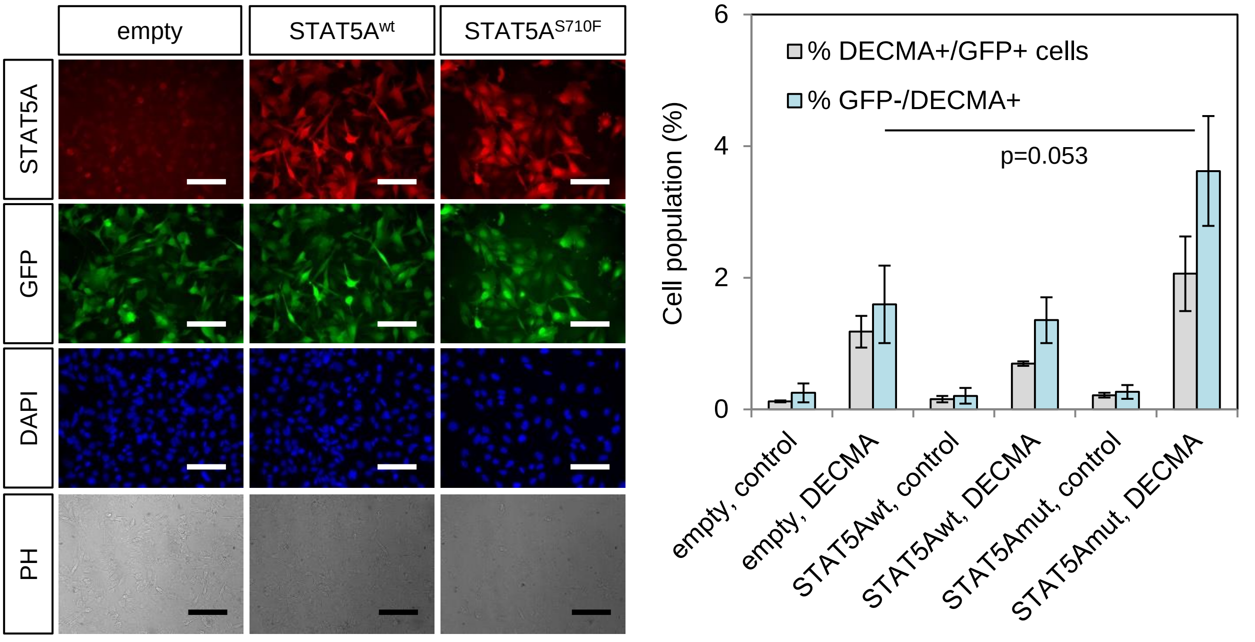

### Supplementary Figure 15

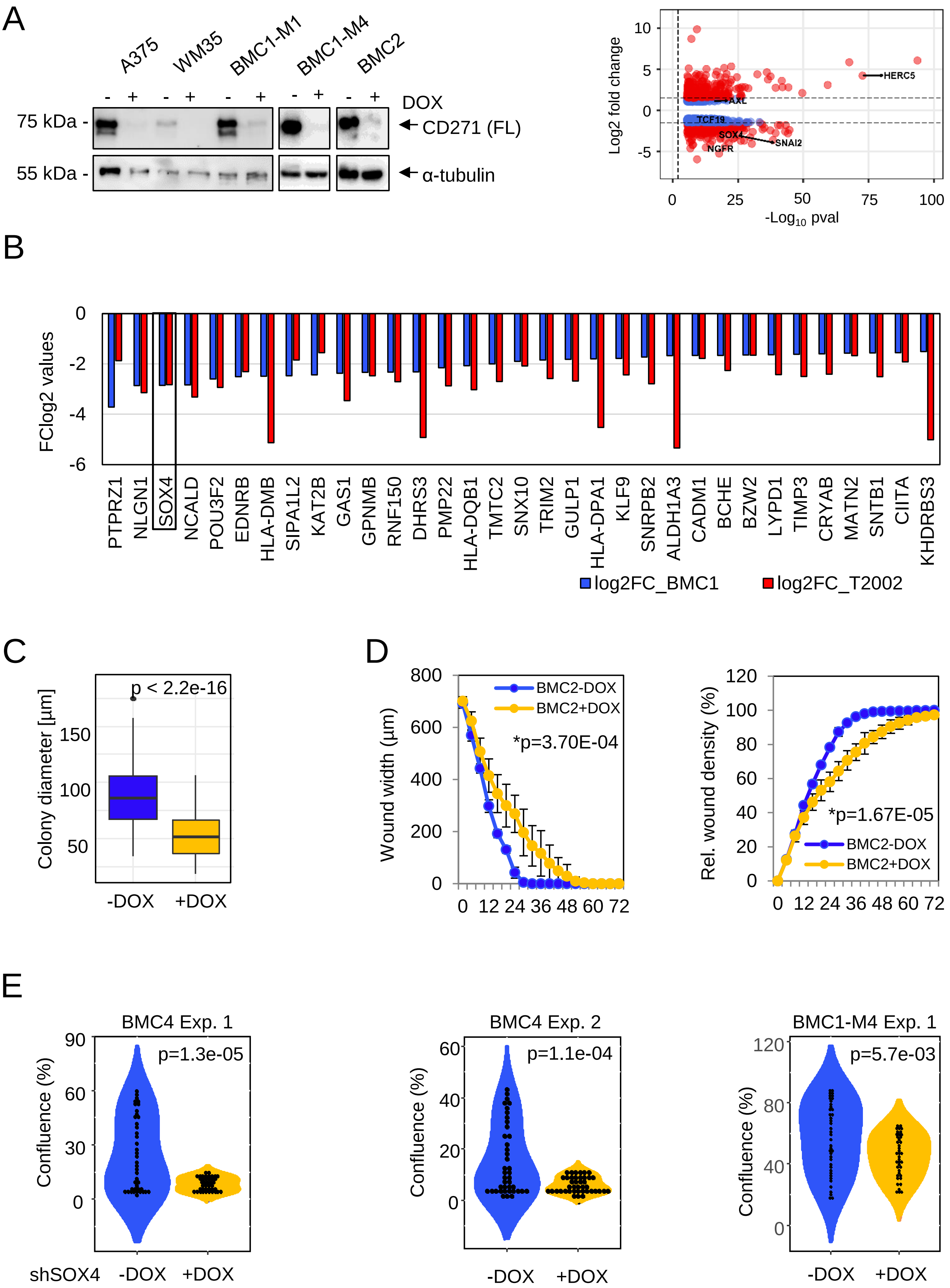
